# “Identification of potential high-risk clones of Carbapenem-Resistant *Pseudomonas aeruginosa* co-producing, KPC-2, VIM-2 and OXA-74 from tertiary hospitals in Metro Manila, Philippines”

**DOI:** 10.1101/2024.06.06.24308581

**Authors:** Sherill Tesalona, Miguel Francisco Abulencia, Maria Ruth Pineda-Cortel, Sylvia A Sapula, Henrietta Venter, Evelina Lagamayo

**Author notes:** Address correspondence to Sherill Tesalona,.

## Abstract

Carbapenem-resistant *Pseudomonas aeruginosa* (CRPA) is an opportunistic human pathogen and a global public health concern due to the limited treatment options available for infections by this pathogen. Identification of antimicrobial resistance (AMR) determinants at an early phase is crucial to administer the appropriate treatment in a timely manner, thereby alleviating the transmission and enhancing the outcomes of patients infected with these pathogens. This study investigated the whole genome sequence (WGS) of 35 CRPA isolates obtained from three hospitals in Metro Manila, Philippines, collected from August 2022 to January 2023. The investigation revealed that a high proportion of isolates obtained from Hospital B and C contained acquired AMR determinants. The traditional multilocus sequence type (MLST) analysis using Pasteur nomenclature in the PubMLST database revealed six different known sequence types (STs), namely ST111, ST175, ST1822, ST357, ST3753 and ST389 and the highest number of unknown STs (27 singletons). Moreover, to delineate the unknown STs identified, the Pasteur nomenclature in the Pathogenwatch database was used, which led to the identification of novel STs. Notably, a novel ST4b1c was found to co-produce the metallo-beta-lactamase (MBL) NDM-7, extended-spectrum beta-lactamases (ESBLs) CTX-M-15 and TEM-2, as well as class D OXA-395. Furthermore, potential high-risk clones represented by ST389 and novel STs (ST95c6, STf555, and STab6a) were identified, all of which co-produced carbapenemases KPC-2, VIM-2, and OXA-74. This study represents the first report in the Philippines on the co-production of ESBL KPC-2, MBL VIM-2 and Class D OXA-74 hosted by potential high-risk clones and novel STs. It underscores the need for continuous surveillance and monitoring of other problematic strains.

## INTRODUCTION

*Pseudomonas aeruginosa* (*P. aeruginosa*) is one of the six leading pathogens for deaths associated with antimicrobial resistance (AMR) and is responsible for an estimated 929,000 deaths attributable to AMR and 3.57 million deaths associated with AMR in 2019 (Murray et al., 2022). *P. aeruginosa* is an opportunistic pathogen (Arzanlou et al., 2017; Raman et al., 2018; Sid Ahmed et al., 2022) that causes serious infections in healthcare environments and among individuals with impaired immune systems (Amsalu et al., 2020; Bogiel et al., 2021). *P. aeruginosa* that is resistant to carbapenem antibiotics, is an emerging and urgent threat (Murray et al., 2022; Willyard, 2017). In 2017, the World Health Organization (WHO) categorized carbapenem-resistant *P. aeruginosa* (CRPA) as “critical,” necessitating the urgent development of new antibiotics (Tacconelli et al., 2018). The emergence and dissemination of the CRPA and the reported “high-risk” clones that are frequently resistant to multiple antibiotics pose an urgent threat to the general welfare of the global population and represents a global risk to public health (Hu et al., 2021; Kocsis et al., 2021; Oliver et al., 2015). The main mechanisms by which *P. aeruginosa* can develop resistance to carbapenems are the following: the production of enzymes that can degrade carbapenem antibiotics, such as the carbapenemases (Amsalu et al., 2021); overexpression of drug efflux pumps, like those in the resistance-nodulation-division (RND) family, which help the cell to export carbapenems (Amsalu et al., 2020); and decreased outer membrane permeability through the downregulation or loss of OprD porins required for carbapenem entry (Li et al., 2012; Shariati et al., 2018; Shu et al., 2017). Carbapenemases are enzymes that can hydrolyze carbapenem antibiotics, rendering them ineffective (Amsalu et al., 2021; Queenan & Bush, 2007). Carbapenemases are classified into two families: serine and metallo-beta-lactamases (MBLs) (Sahuquillo-Arce, 2015). Serine beta-lactamases such as class A (e.g. KPC, PER) and class D beta-lactamases (e.g. OXA-10, OXA-74) that require a serine residue at the active site (Bush & Jacoby, 2010). MBLs are Ambler class B enzymes that require one or two zinc ions for catalytic activity, such as New Delhi metallo-beta-lactamases (NDM) and Verona-Integron metallo-beta-lactamases (VIM) (Bush & Jacoby, 2010). MBLs, unlike serine beta-lactamases, are not inhibited by typical therapeutic inhibitors like clavulanic acid, tazobactam, or sulbactam (Sawa et al., 2020). Their propensity to efficiently hydrolyze practically all carbapenems (Pedroso et al., 2020; Zhao & Hu, 2015) and rapid proliferation compromise existing therapeutic choices (Amsalu et al., 2021). The acquisition of carbapenemases is of great concern as they can hydrolyze most beta-lactam antibiotics (Schäfer et al., 2019) and are often encoded by genes carried on mobile genetic elements such as plasmid, transposon, and insertion sequences, facilitating their transfer to other bacterial species (Yoon & Jeong, 2021).

Carbapenemase production is a rare cause of carbapenem resistance in *P. aeruginosa* isolates in the United States (Karlowsky et al., 2018; Reyes et al., 2023) but is identified in more than 20% of other regions (Gajdács, 2020; Gill et al., 2021; Karlowsky et al., 2018). Studies have reported a prevalence of 18.9% of CRPA in the Asia-Pacific region and a high incidence of carbapenemase-producing *P. aeruginosa* in China and Indonesia (Arowolo et al., 2023; Wang et al., 2021). Furthermore, according to CDC and ECDC reports, the MBL-producing *P. aeruginosa* increased from 4.0% (2021) to 9.0% (2022), and the most commonly identified carbapenemases are VIM, KPC and IMP (CDC, 2019; Nordmann & Poirel, 2019; Reyes et al., 2023; Tacconelli et al., 2017). Several countries in Southeast Asia, including Bangladesh, India, Indonesia, Nepal, Sri Lanka, and Thailand, have observed a worrying trend of increasing drug resistance, while the Philippines was not mentioned in the report (Sihombing et al., 2023). However, a study by Argimon et al., 2020 revealed that in the Philippines, resistance rates to critical pathogen-antibiotic combinations, such as carbapenem-resistant organisms, have steadily increased over the past decade (Argimón et al., 2020). In the annual report of the Antimicrobial Resistance Surveillance Reference Laboratory (ARSRL) of the Philippines, a significant decrease in the resistance rate of *P. aeruginosa* to meropenem (p = 0.0000) from 2021 to 2022 was observed; however, imipenem was never mentioned (Department of Health - RITM, 2022).

In spite of the high incidence of CRPA and the critical need for new antimicrobials against this pathogen, there is a scarcity of research regarding the genetic and molecular epidemiological attributes of CRPA strains in the Philippines. Given that AMR and CRPA, in particular, is a global concern that significantly impact our health and economy, a better understanding of the molecular epidemiology of this critical pathogen is needed to tailor treatments and mitigate the risk for dissemination. Hence, this study reports on the surveillance of CRPA from three hospitals in the Philippines over a six-month period. Whole genome sequencing (WGS) was used to identify resistance genes and to provide insight into the molecular epidemiologic landscape of *P. aeruginosa*.

## RESULTS

### Sample information

A total of 35 CRPA isolates were obtained from a cohort of patients aged 14 to 93 years, 57.0% of which are male. The distribution of CRPA isolates across hospital wards and specimen types was are as follows: 46.0% (n = 16/35) were obtained from the intensive care units (ICU), followed by the medicine department for adults 37.1% (n = 13/35) And pediatrics 2.9% (n = 1/35). The obstetrics and gynecology, and surgery departments contributed 9.0% (n = 3/35) and 6.0% (n = 2/35) of the CRPA isolates analyzed. The 35 CRPA isolates that were investigated were obtained from respiratory specimens (endotracheal aspirate, sputum, and bronchoalveolar lavage), blood, gastric aspirate, wound tissue, ascitic fluid, bone aspirate and catheter drain with the following distributions: 68.0% (n = 24), 11.0% (n = 4) 9.0% (n = 3), 2.9% (n = 1), 2.9% (n = 1), 2.9% (n = 1) and 2.9% (n = 1), respectively (Supplemental File 1).

### The phenotypic AMR profile reveals high proportion of resistance to ciprofloxacin, ceftazidime, and cefepime

The phenotypic analysis exhibited the following incidence of resistance: ceftazidime (CAZ) 60.0% (n = 21/35), cefepime (FEP) 57.1% (n = 20/35), piperacillin-tazobactam (TZP) 48.6% (n = 17/35), amikacin (AMK) 20.0% (n = 7/35), ciprofloxacin (CIP) 62.9% (n = 22/35), and aztreonam (ATM) 38.1% (n = 8/21). Resistance to CIP, CAZ, and FEP was observed from a large proportion of CRPA isolates, 62.9%, 60.0%, and 57.1%, respectively (Table 1). Following the CLSI breakpoints, there were CRPA isolates obtained from Hospital C that showed resistance to Ceftazidime-avibactam (CZA) 63.6%, (n = 7/11), and colistin (COL) 25.0%, (n = 5/20). Interestingly, according to EUCAST, only one (n = 1/20) of the isolates would be COL resistant following the MIC breakpoint of >4 mg/L. However, not all isolates have been subjected to antimicrobial susceptibility test against CZA and COL following the Clinical and Laboratory Standards Institute, 32^nd^ ed, 2022 (CLSI) guidelines (CLSI, 2022) and the institutional (isolation site) protocol. Whereas all the CRPA isolates obtained and investigated are 100.0% IPM resistant, only 82.9% (n=29/35) are tested MEM-resistant (Table 1 and 2). The phenotypic AMR is shown in Supplemental File 2.

**Table 1:** MIC distribution for 35 CRPA isolates obtained from three tertiary hospitals in Metro Manila, from August 2022 to January 2023 following the CLS1, 2022 guidelines.

| Antibiotics | No. of isolates at an MIC (mg/L) of: |  |  |  |  |  |  |  |  |  |  | Breakpoint(s)<sup>b</sup> | R (%) |
| --- | --- | --- | --- | --- | --- | --- | --- | --- | --- | --- | --- | --- | --- |
|  | 0.125 | 0.250 | 0.5 | 1 | 2 | 4 | 8 | 16 | 32 | 64 | 128 |  |  |
| IPM<sup>d, e</sup> |  |  |  |  |  |  |  | 35<sup>+, a</sup> |  |  |  | ≤2/≥8 | 100.0 |
| MEM<sup>d</sup> |  | 1 * |  |  |  | 5 | 3<sup>+, a</sup> | 1, 25<sup>+, a</sup> |  |  |  | ≤2/≥8 | 82.9 |
| CAZ<sup>d</sup> |  |  |  |  |  | 11 |  | 3, 7<sup>+</sup> | 1<sup>+, a</sup> | 13<sup>+, a</sup> |  | ≤8/≥32 | 60.0 |
| FEP<sup>d</sup> |  |  |  | 5 * | 3 |  | 3 | 4, 6<sup>+</sup> | 1<sup>a</sup> | 13<sup>+, a</sup> |  | ≤8/≥32 | 57.1 |
| CZA<sup>e</sup> |  |  |  |  | 1 | 3 | 7<sup>+</sup> |  |  |  |  | (≤8/4)/(≥16/4) | 63.6 |
| TZP<sup>d</sup> |  |  |  |  |  | 4 * | 1 | 4 | 9 | 12<sup>+</sup> | 5<sup>+, a</sup> | (≤16/4)/(≥128/4) | 48.6 |
| COL<sup>e</sup> |  |  |  | 9 * | 6 | 4<sup>a</sup> |  | 1<sup>+, a</sup> |  |  |  | ≤2/≥4 | 25.0 |
| CIP<sup>d</sup> | 1* | 4* | 6* | 2 | 10<sup>+, a</sup> | 11<sup>+, a</sup> |  |  |  | 1<sup>+, a</sup> |  | ≤0.5/≥2 | 62.9 |
| AMK<sup>d</sup> |  |  |  |  | 10 * | 3 | 9 * | 3 | 3 | 7<sup>+, a</sup> |  | ≤16/≥64 | 20.0 |
|  | No. of isolates at zone diameter (mm) <sup>b</sup> of: |  |  |  |  |  |  |  |  |  |  | Breakpoint(s)<sup>c</sup> |  |
|  | 12 | 13 | 14 | 15 | 16 | 17 | 18 | 19 | 20 | 21 | 22 |  |  |
| ATM<sup>f</sup> | 1<sup>a</sup> |  |  | 7<sup>a</sup> | 1 | 1 |  |  | 4 |  | 7 | ≥22/≤15 | 38.1 |

CRPA, carbapenem-resistant *P. aeruginosa*; AMR, Antimicrobial resistant; IPM, imipenem; MEM, meropenem; CAZ, ceftazidime; FEP, cefepime; CZA, ceftazidime-avibactam; TZP, piperacillin-tazobactam; AMK, amikacin; CIP, ciprofloxacin; COL, colistin; and ATM, aztreonam.
<sup>a</sup> indicates data for resistant strains as per CLSI 2022; \* MIC ≤ the indicated value; + MIC > the indicated value; <sup>b</sup> Breakpoint concentrations - expressed as "susceptibility (mg/L)/resistance (mg/L)" and <sup>c</sup> Breakpoint concentrations - expressed as "zone of inhibition in mm" - are presented according to CLSI 2022; <sup>d</sup> MIC detected using Vitek®2 system (bioMérieux); <sup>e</sup> MIC detected using Thermo Scientific™ Sensititre™; and <sup>f</sup> MIC detected using Kirby Bauer technique.

### Differences in the prevalence rate and level of resistance to third (CAZ) and fourth-generation cephalosporin (FEP), and fluoroquinolone (CIP) among the three hospitals

The 35 CRPA isolates investigated are relatively not equally distributed among the three isolation sites: (Hospital A, n = 8/35), Hospital B, n = 16/35, and Hospital C, n = 11/35), 22.9%, 45.7%, and 31.4%, respectively. Hospital B has the highest number of CRPA isolates investigated in this study. Analysis revealed that 62.5 % (n =10/16) of CRPA isolates obtained from Hospital B showed resistance to CAZ, FEP, and CIP, while 72.7% (n = 8/11) of CRPA isolates obtained from Hospital C showed resistance to both CAZ and FEP and a high proportion of CIP resistance, 90.9% (n = 10/11) (FIG 1A). Moreover, low proportion of resistance against CAZ, FEP, and CIP was observed in CRPA isolates obtained from Hospital A. Only 37.5% (n = 3/8) CRPA isolates from Hospital A showed resistance to the third-generation cephalosporin CAZ, and 25.0% (n = 2/8) showed resistance against fourth-generation cephalosporin FEP, and fluoroquinolone (CIP). The incidence rate and resistance level against CZA and COL cannot be compared from the three isolation sites because the institutions have different protocols for testing multidrug-resistant organisms (MDROs) against these antibiotics. Of the three isolation sites, only Hospital C has automatically included testing CZA and COL against MDROs and reported following the cascade reporting rules and upon the physician’s request. Out of the 11 isolates tested from Hospital C, 63.6% (n = 7/11) expressed resistance against CZA (MIC >8 mg/L) and 18.2% (n = 2/11) expressed resistance against COL (MIC ≥4 mg/L) (FIG 1A, Tables 1 and 2 and Supplemental File 2).

**FIG 1A.**
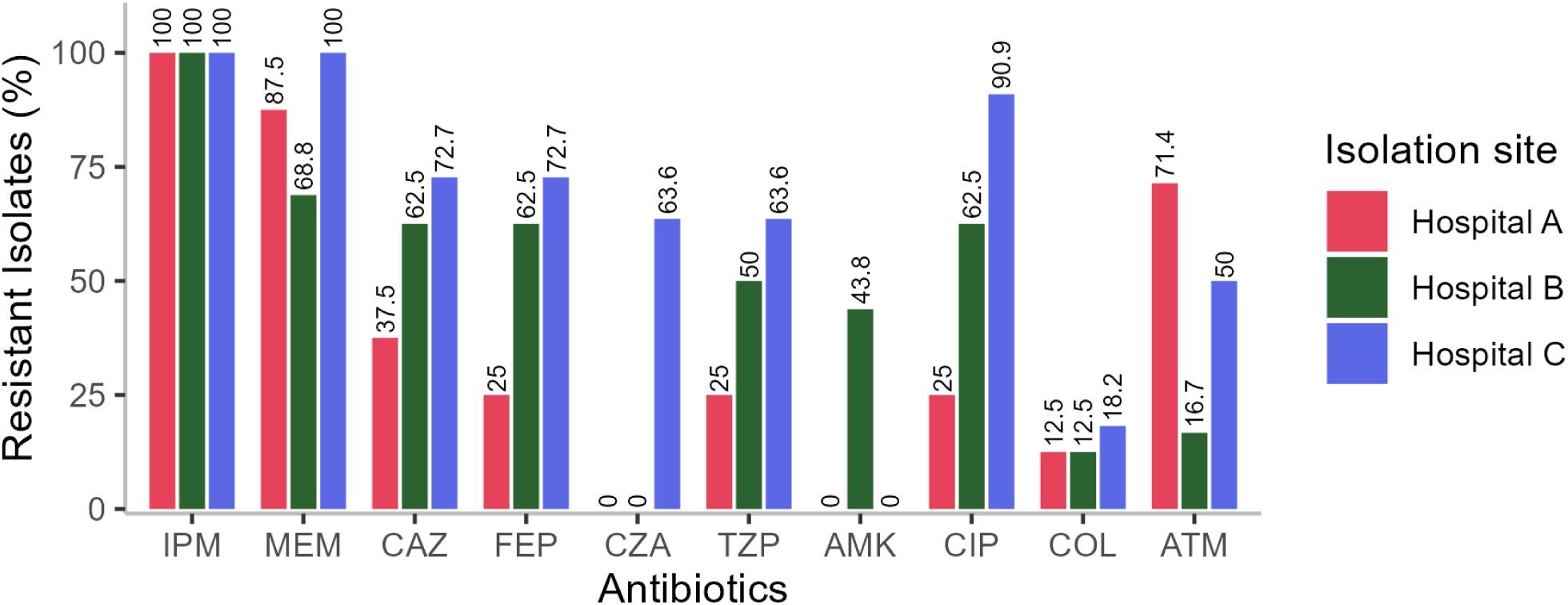
Percentage of resistant isolates in the three sampling sites. IPM, imipenem; MEM, meropenem; CAZ, ceftazidime; FEP, cefepime; CZA; ceftazidime-avibactam; TZP, piperacillin-tazobactam; AMK, amikacin; CIP, ciprofloxacin; COL, colistin; TOB, tobramycin; and ATM, aztreonam.

**Table 2.** CLSI breakpoints, and minimum inhibitory concentrations (MICs) for antibiotics used in this study and sample type information of 35 CRPA isolates.

|  |  | MIC (mg/L) |  |  |  |  |  |  |  | Zone Diameter Breakpoints (mm) |  |
| --- | --- | --- | --- | --- | --- | --- | --- | --- | --- | --- | --- |
|  |  | IPM<sup>a</sup> | MEM<sup>a</sup> | CAZ<sup>a</sup> | FEP<sup>a</sup> | CZA<sup>b</sup> | TZP<sup>a</sup> | AMK<sup>a</sup> | CIP<sup>a</sup> | COL<sup>b</sup> | ATM<sup>c</sup> |
| Isolate | Specimen Type | ≥8 | ≥8 | ≥32 | ≥32 | ≥16 | ≥128 | ≥64 | ≥2 | ≥4 | ≤15 |
| P01T | BAL | >16 | >16 | 16 | 16 | N/T | 32 | <2 | 1 | >16 | 15 |
| P02T | Blood | >16 | >16 | 4 | 2 | N/T | 16 | <2 | 2 | N/T | N/T |
| P03T | ETA | >16 | 4 | >32 | 8 | N/T | 32 | <2 | <0.25 | N/T | 20 |
| P04T | Blood | >16 | >16 | 4 | 2 | N/T | 16 | <2 | 2 | N/T | 20 |
| P05T | BAL | >16 | >16 | 16 | 16 | N/T | 32 | 16 | 1 | N/T | 15 |
| P06T | BA | >16 | >16 | >64 | >64 | N/T | >64 | <2 | <0.25 | N/T | 12 |
| P07T | Sputum | >16 | >16 | >64 | >64 | N/T | >64 | <2 | 0.5 | N/T | 15 |
| P08T | ETA | >16 | >16 | 4 | 8 | N/T | 32 | <2 | <0.25 | N/T | 15 |
| P01L | ETA | >16 | >8 | 4 | ≤1 | N/T | 32 | 8 | 0.5 | N/T | 16 |
| P02L | Blood | >16 | 4 | 4 | ≤1 | N/T | <4 | <8 | 0.5 | N/T | 22 |
| P03L | Blood | >16 | >8 | >64 | >64 | N/T | 32 | 32 | >4 | N/T | 22 |
| P04L | Sputum | >16 | >8 | >64 | >64 | N/T | >128 | 4 | <0.125 | N/T | 22 |
| P05L | ETA | >16 | >16 | >64 | >64 | N/T | >64 | >64 | >64 | N/T | 15 |
| P06L | Wound Tissue | >16 | >16 | >64 | >64 | N/T | 32 | <2 | >4 | 4 | N/T |
| P07L | ETA | >16 | 4 | 4 | ≤1 | N/T | <4 | 4 | >4 | N/T | 15 |
| P08L | Catheter Drain | >16 | 4 | 4 | ≤1 | N/T | <4 | 4 | <0.5 | N/T | 22 |
| P09L | Gastric Aspirate | >16 | ≤ 0.25 | 4 | ≤1 | N/T | <4 | <2 | <0.5 | 2 | 22 |
| P10L | Gastric Aspirate | >16 | 4 | 4 | 2 | N/T | 32 | <2 | <0.5 | 2 | N/T |
| P11L | ETA | >16 | >16 | >64 | >64 | N/T | >128 | >64 | >4 | 4 | 20 |
| P12L | ETA | >16 | >16 | >64 | >64 | N/T | >64 | >64 | >4 | N/T | 17 |
| P13L | ETA | >16 | >16 | >64 | >64 | N/T | >128 | >64 | >4 | 2 | 20 |
| P14L | ETA | >16 | >16 | >64 | >64 | N/T | >128 | >64 | >4 | 2 | N/T |
| P15L | Ascitic fluid | >16 | >16 | >64 | >64 | N/T | >64 | >64 | >4 | 2 | 22 |
| P16L | ETA | >16 | >16 | >64 | >64 | N/T | >128 | >64 | >4 | 2 | N/T |
| P01C | Sputum | >16 | >16 | 4 | 8 | 4 | 8 | <8 | >2 | <1 | N/T |
| P02C | Sputum | >16 | >16 | >16 | >16 | >8* | >64 | <8 | >2 | <1 | N/T |
| P03C | Sputum | >16 | >16 | >16 | 16 | 2 | >64 | <8 | <0.125 | <1 | N/T |
| P05C | ETA | >16 | >16 | 4 | 32 | >8* | 16 | 8 | >4 | 4 | 15 |
| P06C | ETA | >16 | >16 | >64 | >64 | >8* | 32 | <2 | >4 | 4 | N/T |
| P07C | Sputum | >16 | >16 | >16 | >16 | >8* | >64 | <8 | >2 | <1 | 22 |
| P08C | Sputum | >16 | >16 | 16 | 16 | 4 | 16 | 32 | >2 | <1 | N/T |
| P09C | Gastric Aspirate | >16 | >16 | >16 | >16 | >8* | >64 | <8 | >2 | <1 | N/T |
| P10C | Sputum | >16 | 16 | >16 | >16 | >8* | >64 | 16 | >2 | <1 | N/T |
| P11C | Sputum | >16 | >16 | >16 | >16 | 4 | >64 | 32 | >2 | <1 | N/T |
| P12C | Sputum | >16 | >16 | >16 | >16 | >8* | >64 | 16 | >2 | <1 | N/T |

MIC, minimum inhibitory concentration; IPM, imipenem; MEM, meropenem; CAZ, ceftazidime; FEP, cefepime; CZA, ceftazidime-avibactam; TZP, piperacillin-tazobactam; AMK, amikacin; CIP, ciprofloxacin; COL, colistin; TOB, tobramycin; ATM, aztreonam; OprD, outer membrane porin D; AMR, Antimicrobial resistance; BAL, bronchoalveolar lavage; BA, bone marrow aspirate; ETA, endotracheal aspirate; And CRPA, carbapenem-resistant *P. aeruginosa*.
\*, with >8 mg/L MIC reported as R --resistant and an "Intermediate category" is not provided by the CLSI 2022 guidelines. N/T, Not Tested -CLSI guidelines recommend the Antibiotics may warrant primary testing, but their reporting may be selective, such as when the organism is resistant to agents of the same antimicrobial class or for infection control purposes
<sup>a</sup> MIC detected using Vitek®2 system (bioMérieux); <sup>b</sup> MIC detected using Thermo Scientific™ Sensititre™; <sup>c</sup> MIC detected using Kirby Bauer technique MIC breakpoints were based on CLSI 2022

A large proportion of CRPA isolates obtained from Hospital B and C showed resistance against CAZ and FEP (62.5% and 72.7%), in contrast to the incidence of CAZ and FEP resistance in Hospital A, (37.5% And 25.0%) (FIG 1A). The same observation could be made for CIP resistance. MICs revealed moderate-level CAZ and FEP resistance (MICs >64 mg/L) against isolates obtained from Hospital A and B in contrast to Hospital C (FIG 1A).

The MIC values of CRPA isolates showed resistance to multiple classes of antibiotics. All isolates assessed in this study were resistant to at least one antibiotic (FIG 1B). The MDR phenotype was observed in CRPA isolates obtained from Hospital C, 100.0% (n = 11/11), Hospital B, 75.0% (n = 12/16) and Hospital A, 87.5% (n = 7/8) and showed resistance to at least one antibiotics in three or more antimicrobial class-tested (FIG 1B, 1C, and Supplemental File 2). Out of the 16 CRPA isolates investigated from Hospital B, 75.0% (n = 12/16) are multidrug-resistant (MDR). Of these, 68.75% (n = 11/16) expressed resistance against three or more groups of antibiotics, while 18.2% (n = 2/11) expressed resistance to eight antibiotics tested (FIG 1B and FIG 1C).

**FIG 1B.**
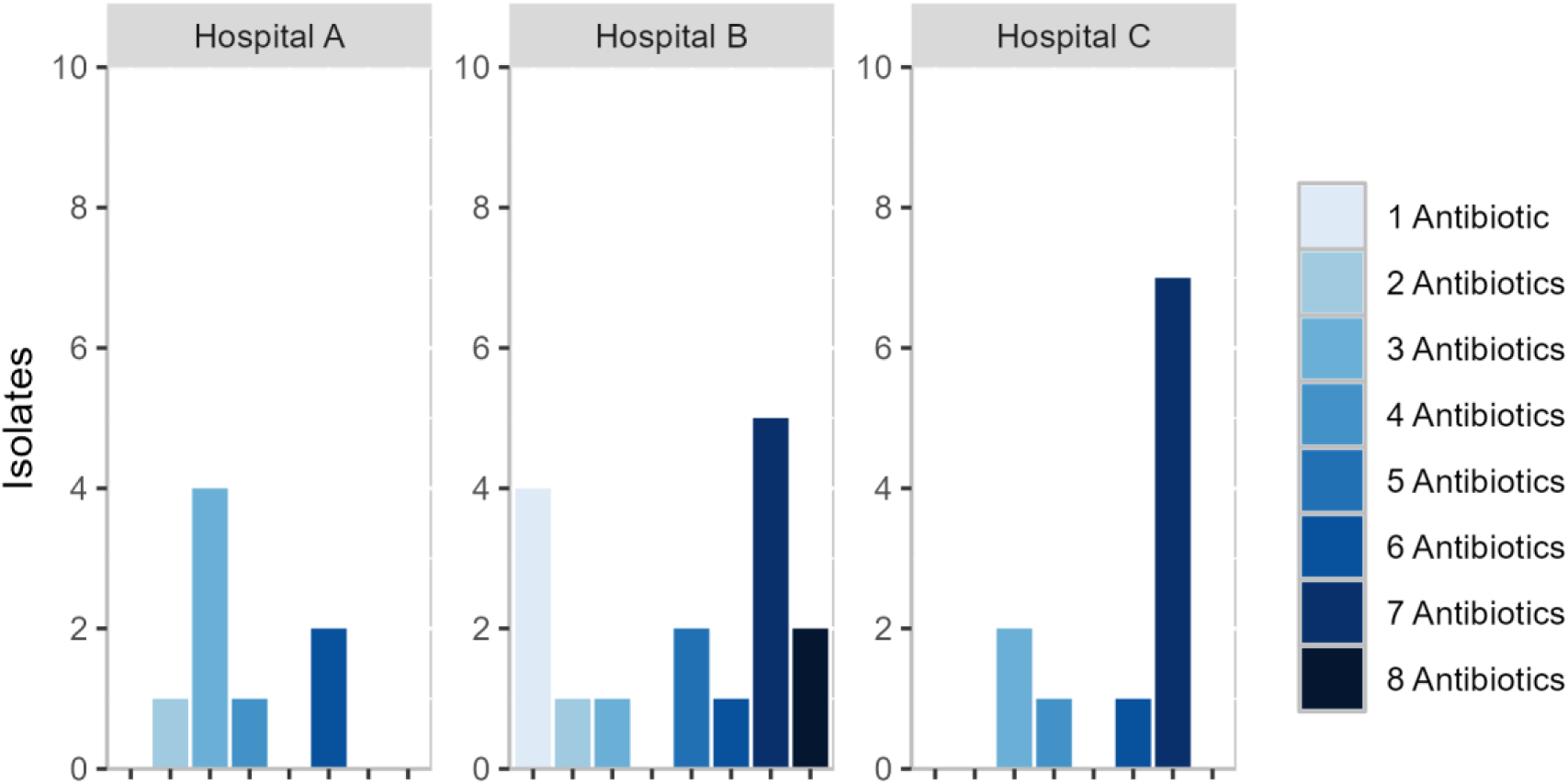
Prevalence of CRPA isolates (n = 35) per facility and the number of antibiotics (n = 8) each isolate was resistant against. The CRPA isolates from Hospitals B and C showed resistance against a greater number of different antibiotics.

**FIG 1C.**
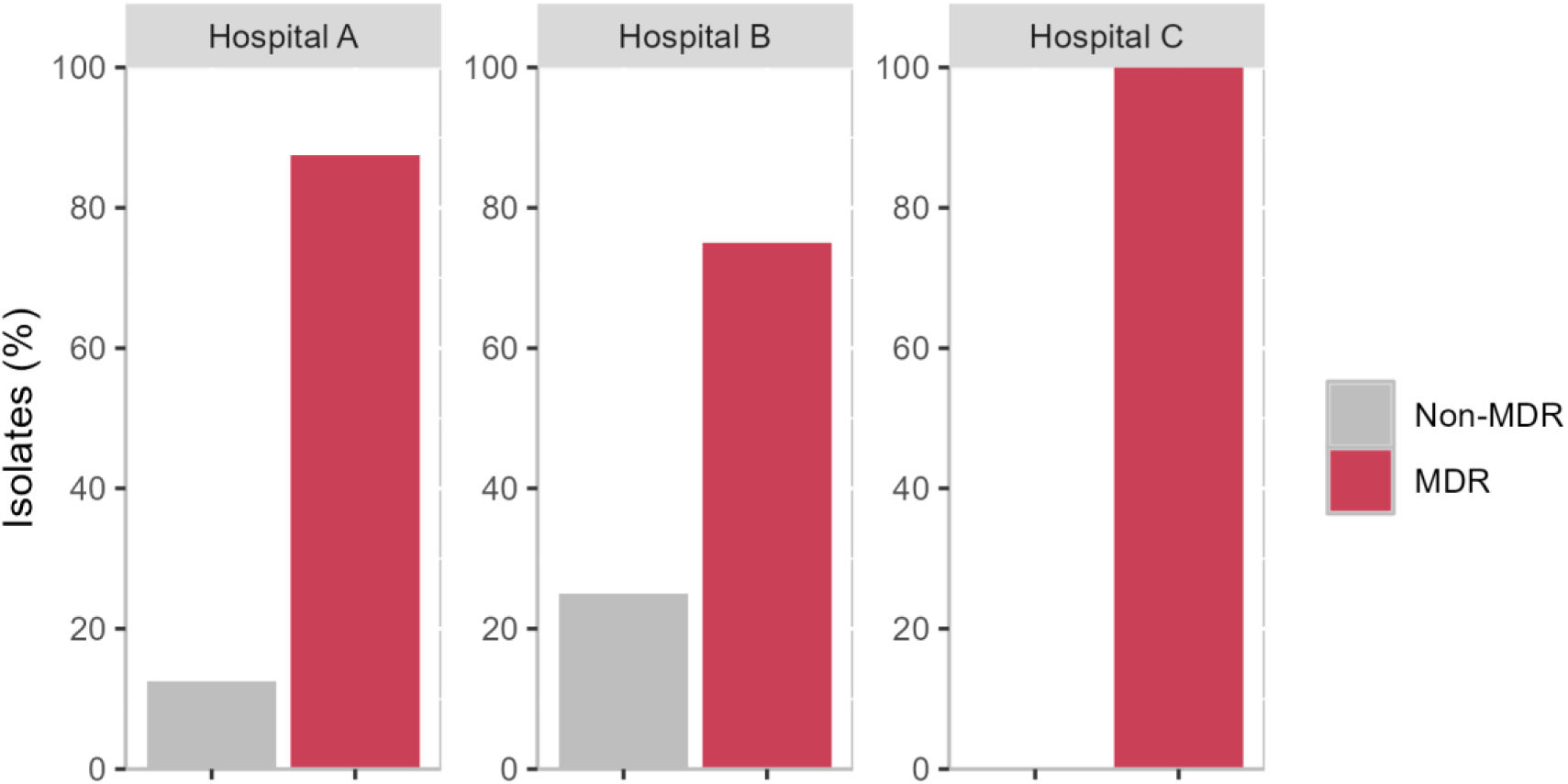
Number of MDR and non-MDR CRPA isolates per isolation site. Hospital A, 87.5% (n = 7/8); Hospital B, 75.0% (n = 12/16); and Hospital C, 100.0% (n = 11/11).

#### Genotyping using MLST analysis for 35 CRPA isolates

Of the 36 CRPA isolates subjected to whole genome sequencing, 35 CRPA isolates, eight (22.9%) from Hospital A, 16 (45.7%) from Hospital B and 11 (31.4%) from Hospital C, met the quality control (QC) standards.

The genome validation process. (1) Sequence Read Quality: The raw sequences underwent read trimming and quality assessment using Fastp (Chen, et al., 2018), ensuring a high Phred score of 30 (Q30) for the sequence reads. (2) Genus-Level Taxonomic Classification: Using the Kraken2 Minikraken database (Wood et al., 2019; Wood & Salzberg, 2014), at least 80% of the reads that passed the filter belonged to the Pseudomonas genus (Vivo et al., 2022). This high percentage of genus-specific reads confirms the predominance of the target organisms in the sequenced samples, minimizing the presence of contaminating sequences from other genera. (3) Genome Coverage: The number of reads provided at least 30x read coverage for *P. aeruginosa* genome. The read coverage of 30x is generally considered sufficient for accurate genome assembly and variant calling (Liu et al., 2022; Seah et al., 2023) . The genome assembly and variant calling were performed in the main workflow of the EpiTomas bioinformatics pipeline. The high coverage ensures a comprehensive representation of the genomes, allowing for reliable downstream analyses such as resistance gene identification and strain typing. (4) Sequencing Depth: The total number of reads analyzed for each *P. aeruginosa* genome exceeded two million. The high sequencing depth ensures that the genomes are well-represented and that the data is suitable for downstream analyses, such as comparative genomics and identifying genetic determinants associated with antimicrobial resistance. Additionally, the GC content was also checked. The included *P. aeruginosa* genomes have a GC content ranging from 64.9 to 67.2%. These GC content ranges are consistent with the expected values for these species, further confirming the accurate identification of the target organisms.

#### Sequence alignment against the RefGenome PA01

After pre-processing, sequence reads underwent alignment against PA01 NC_002516.2 as the reference genome for *P. aeruginosa* using snippy and subsequently determined the alignment coverage using mosdepth (Pedersen & Quinlan, 2018), followed by variant calling. Additionally, the predicted effect and annotation of the called variants were performed using SnpEff (Cingolani et al., 2012). The PA01, as the chosen reference genome for *P. aeruginosa*, with RefSeq accession NC_002516.2; the genome size is 6.2 Mbp, total genes of 5,697, protein-coding genes of 5,572 and with GC content of 66.5%, and available at https://www.ncbi.nlm.nih.gov/datasets/genome/GCF_000006765.1/.

By meeting these quality control standards, the included samples are suitable for downstream analyses, such as MGEs and resistance gene identification, strain typing, and mutation identification, which will provide valuable insights into the epidemiology and resistance mechanisms of *P. aeruginosa*.

Traditional MLST and cgMLST were used here to investigate the high-risk clones and genomic epidemiology using the consensus sequence of the 35 CRPA isolates investigated in this study.

#### A. Traditional MLST analysis

The traditional MLST analysis using Pasteur nomenclature in the PubMLST database (Jolley et al., 2018) revealed the highest number of unknown STS (27 singletons) and six different known STs among the 35 sequenced CRPA isolates (FIG 2A). The international high-risk clone ST111 was represented by one isolate, 6.25% (n = 1/16) obtained from Hospital B. Hospital B was also represented by ST1822 (n = 1/16, 6.25%), ST357 (n = 1/16, 6.25%), ST389 (n = 1/16, 6.25%), and unknown STs represented the largest ST (n = 12/16, 75%) of the CRPA isolates obtained from this isolation site. Moreover, another global high-risk clone, ST175, was represented by the two isolates, 18.18% (n = 2/11) obtained from Hospital C, and unknown STs represented the largest ST (n = 9/11, 81.82%) of the CRPA isolates obtained from this isolation site. Hospital A was represented by ST3753, 25.0% (n = 2/8), and predominantly unknown STs, 75.0% (n = 6/8) (FIG 2A).

**FIG 2A.**
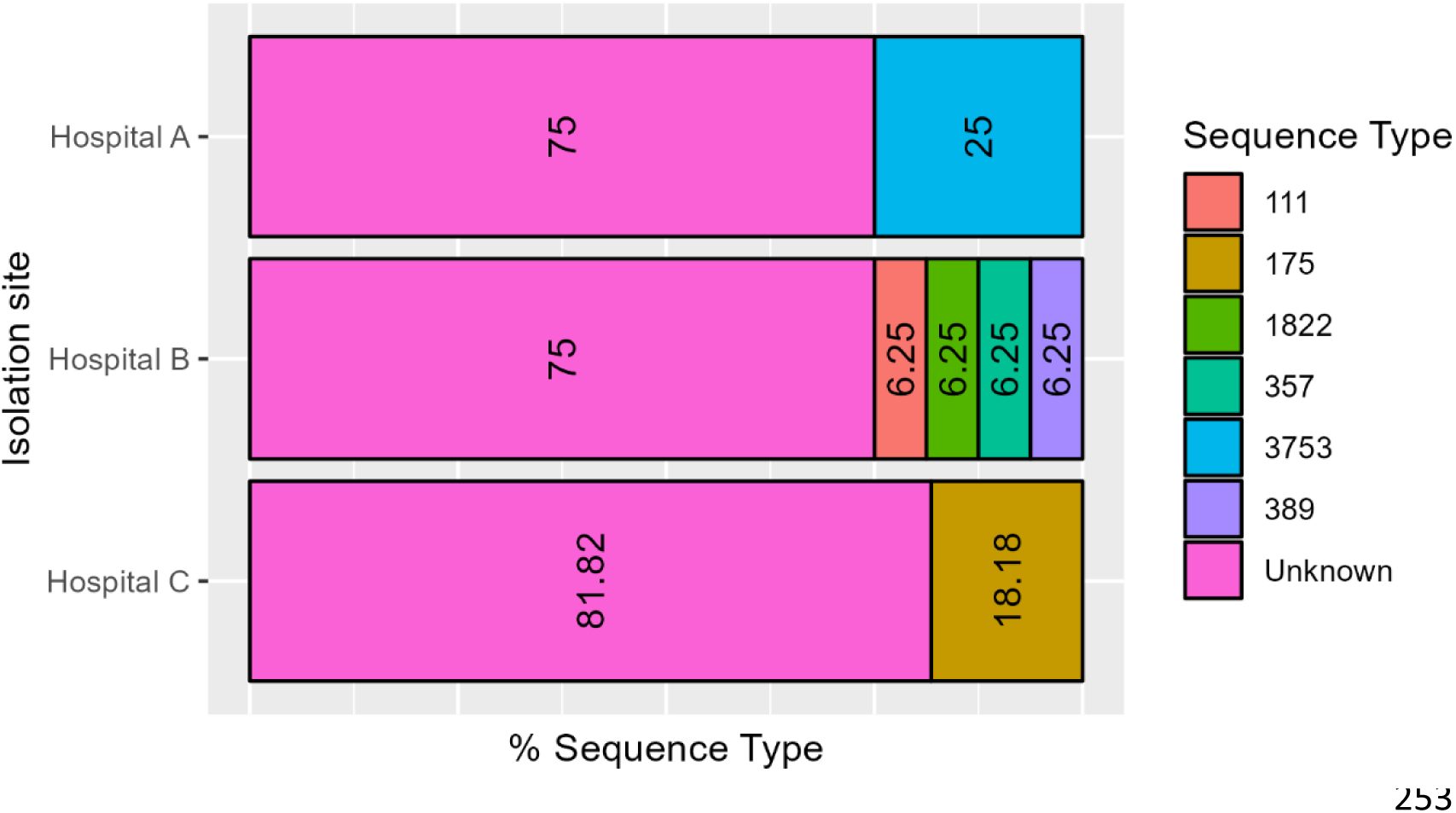
Distribution and proportion of STs of CRPA isolates per hospital using the Pasteur nomenclature in the PubMLST database (Jolley et al., 2018). The number inside the graph denotes the percentage of ST identified. The high percentage of unknown ST was identified and represented by the CRPA obtained from the three isolation sites. The right side denotes the different STs identified.

However, the traditional MLST analysis using Pasteur nomenclature in the Pathogenwatch database (https://cgps.gitbook.io/pathogenwatch/technical-descriptions/typing-methods/mlst) revealed that 77.1% (n = 27/35) of CRPA isolates, represented by singleton unknown STs possessing novel allele/s represented by CRPA isolate ST with an asterisk * (FIG 2B And Supplemental File 3). An asterisk *represents novel ST with a specific allele that is unavailable or unknown, and ∼, an almost similar allele https://cgps.gitbook.io/pathogenwatch/technical-descriptions/typing-methods/mlst (Fig 2B). Hospitals A, B And C were represented by the following proportion of unknown/novel STs: 75.0% (n = 6/8), 75.0% (n = 12/16), and 81.82% (n = 9/11), respectively (FIG 2A). In summary, 55.6% (n = 15/27) of singleton new STs possessed new alleles, while 25.9% (n = 7/27) possessed multi-allelic (7,321) at *trpE* locus. The multi-allelic CRPA isolates were represented by P08L, P11L, P12L, P15L, and P16L CRPA isolates, obtained from Hospital B, and P10C and P12C CRPA isolates obtained from Hospital C (Supplemental File 3).

**FIG 2B.**
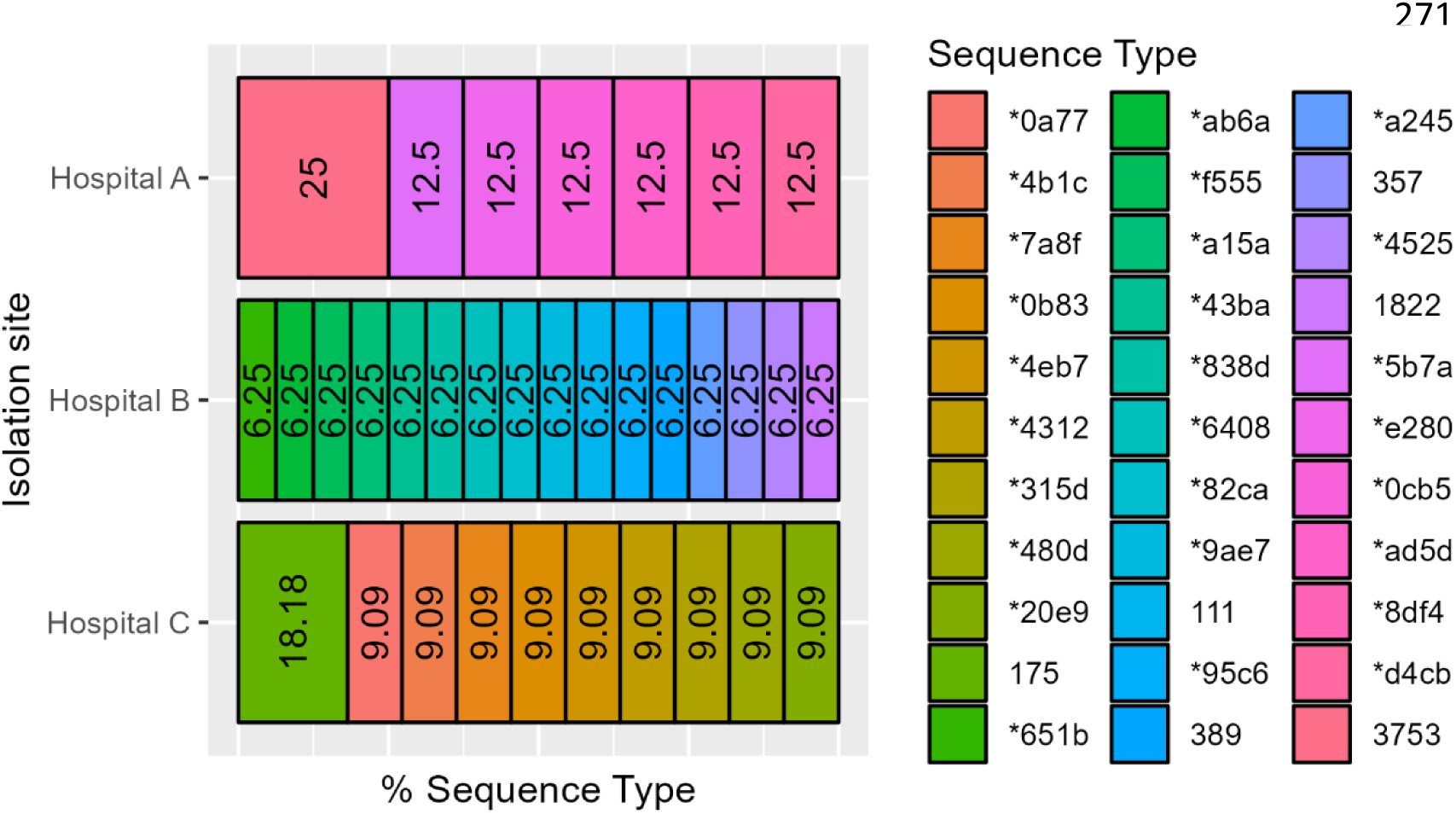
Distribution and proportion of sequence type (ST) CRPA isolate per hospital using Pasteur nomenclature in the Pathogenwatch database (https://cgps.gitbook.io/pathogenwatch/technicaldescriptions/typing-methods/mlst). The right side denotes the different STs identified. Asterisk *represents novel ST with a specific allele that is unavailable or unknown. The number inside the graph denotes the percentage of ST identified. Unique or singleton novel ST were identified and represented by the CRPA isolates obtained from the three isolation sites.

Based on the traditional MLST analysis, the minimum spanning was constructed. The following CRPA isolates represented the index case of each isolation site: Hospital A, P01T, Hospital B, P01L, and Hospital C, P01C. In this manuscript, the term index case refers to the first CRPA isolates in August 2022, which marked the beginning of the isolate collection from the three isolation sites. Clustering from the index cases of CRPA infections from each isolation site was not observed (FIG 3A). The index cases from each isolation site are the following: P01T, P01L and P01C.

**FIG 3A.**
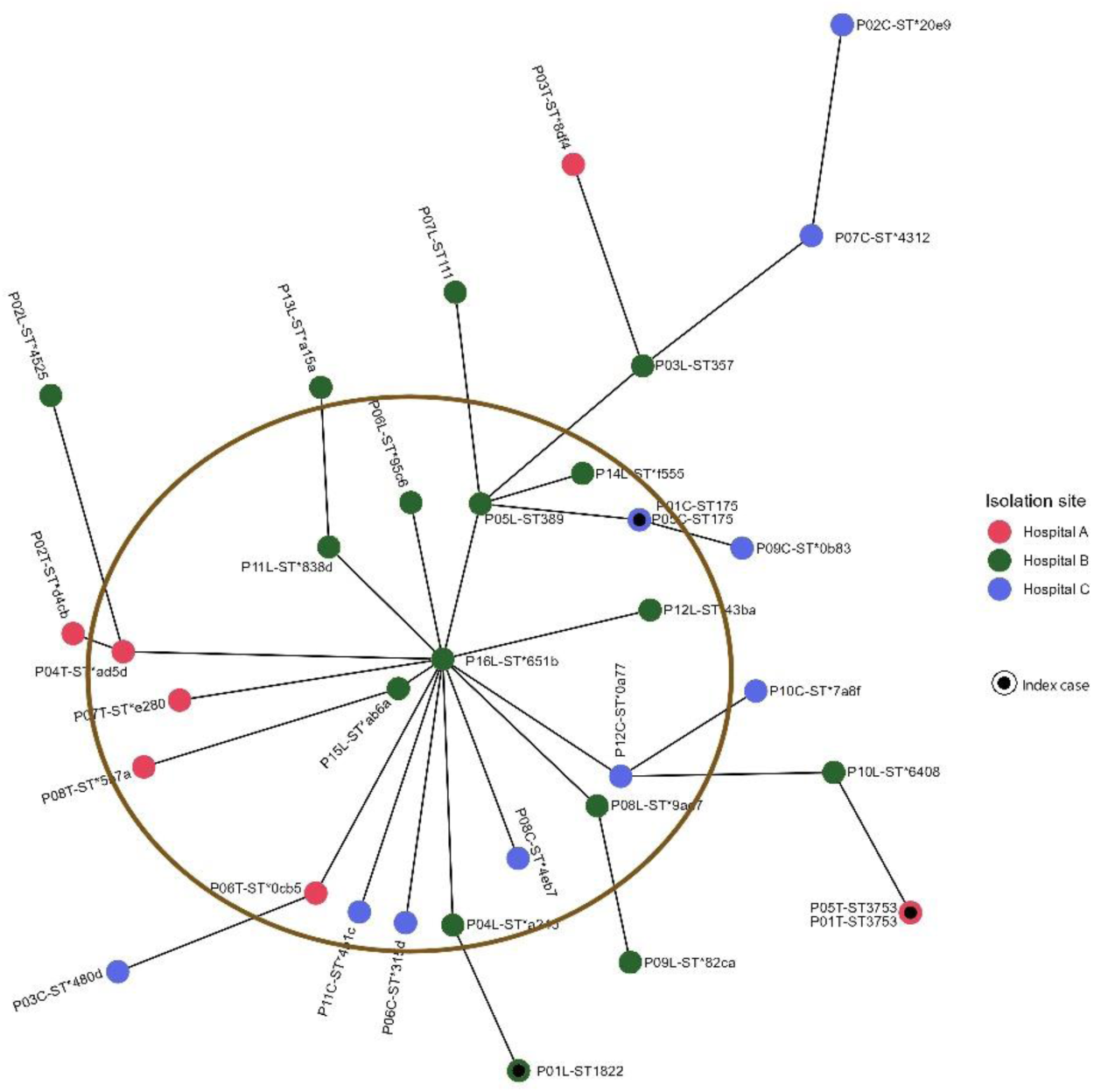
Genotyping of 35 CRPA isolates based on traditional MLST scheme utilizing Pasteur nomenclature in the Pathogenwatch and the PubMLST database (Jolley et al., 2018). The circles are named according to the isolates and colored according to the isolation sites. Each circle includes the ST. The inner black circles represent the index case. In this manuscript, the index case is the first case of CRPA infections in August 2022, which marked the beginning of the isolate collection from the three isolation sites. The circles labeled with more than one sample represent that these samples exhibited identical ST. The major cluster involving the P16L is indicated in a circle shown in FIG 3A.

The traditional MLST used the schema of seven housekeeping genes (*acsA*, *aroE*, *guaA*, *mutL*, *nuoD*, *ppsA*, and *trpE*), which led to the typing and identification of two global high-risk clones: ST111 represented by P07L CRPA isolate obtained from Hospital B, and ST175 represented by P01C and P05C CRPA isolates obtained from Hospital C. However, one major cluster was observed involving P16L CRPA isolate represented by ST*651b, which was a sample obtained from Hospital B and was linked to 13 isolates obtained from three different hospitals represented by new STs and one isolate obtained from Hospital B represented by ST389. These CRPA isolates clustered to P16L, which was an isolate obtained from Hospital B, were observed to have shared similar alleles in two or more loci. However, the allele profiling is based or limited only to seven loci or seven *P. aeruginosa* housekeeping genes. Furthermore, the P05C and P01C isolates obtained from Hospital C were both represented as ST175, having allele differences at locus *trpE* (19 and 134) when compared to P09C isolate, and allele differences at loci *acsA* (28, 17), *nuoD* (3, 1) and *trpE* (19, 3) when compared to P05L isolate that was obtained from Hospital B. Additionally, The P01L CRPA isolate, which was the index case of Hospital B, is the single ST1822 identified. Finally, two CRPA isolates, namely P05T and P01T, which were obtained from Hospital A, were both represented as ST3753. The genotyping of the 35 CRPA isolates using traditional MLST unveiled the ubiquity of *P. aeruginosa* and required the application of cgMLST to delineate the sporadic cases and provide high-resolution genetic distinction among 35 CRPA isolates obtained from the three isolation sites. The details of the allelic profile between 35 CRPA isolate and the wild-type PA01 are shown in Supplemental File 3.

#### B. cgMLST Analysis

We reanalyzed the consensus sequence of 35 CRPA isolates employing the high-resolution cgMLST schema (Tönnies et al., 2021). The cgMLST analysis for *P. aeruginosa* is not yet hosted by the Pathogenwatch database (https://Pathogenwatch/). Hence, the cgMLST.org database (de Sales et al., 2020) was used together with the pyMLST tool, which uses the wild-type PA01 as the seed genome (Biguenet et al., 2023). This was done due to the highly ubiquitous characteristics of the 35 CRPA isolates and the involvement of multiple sequence types, both known and new STs identified by the traditional MLST analysis.

Supplemental File four depicts the allelic profile of 35 CRPA isolates based on the cgMLST schema, which analyses up to 3,867 target genes and up to 1,265,835 alleles to increase the resolution in gene characterization. The header corresponds to the loci name established by the cgMLST schema of Tonnies et al., 2021. Each number below each locus corresponds to the allele profile of each CRPA genome (e.g. WTPA01 has allele 1 at locus PA3729 while P01C has allele 3,868 at the same locus). Whereas, Supplemental File five depicts the number of allele differences or gene distinctions of each CRPA isolate based on 3,867 target genes, far from seven target genes utilized in the traditional MLST analysis (e.g. Out of 3,867 target genes used by the cgMLST schema, WTPA01 is distinct from P01C for up to 2,828 target genes, whereas, P01L is distinct from P01C for up to 3,631 target genes).

A minimum spanning tree was constructed based on the cgMLST schema. Clustering was not observed from the index cases of CRPA infections, namely P01T, P01L, and P01C isolates, in three isolation sites, namely Hospital A, B, and C, based on the number of allelic differences utilizing up to 3,867 target genes and 1,265,835 alleles (FIG 3B). However, a major cluster involving the P06L is indicated in a circle shown in FIG 3B. P06L CRPA isoate was obtained from Hospital B and linked to 17 CRPA isolates obtained from three isolation sites having the allelic distinction of 1,664 to 3,551 alleles (FIG 3B). P05L CRPA isolate obtained from the same hospital (Hospital B) had the nearest genetic distinction when compared with P06L isolate, having 1,664 allele differences, while P03T isolate obtained from Hospital A has the farthest or highest genetic distinction corresponding to 3,551 allelic differences from P06L isolate (Supplemental File 5 and FIG 3B). These findings showed that the sequence diversity of *P. aeruginosa* was high among the 35 isolates investigated.

**FIG 3B.**
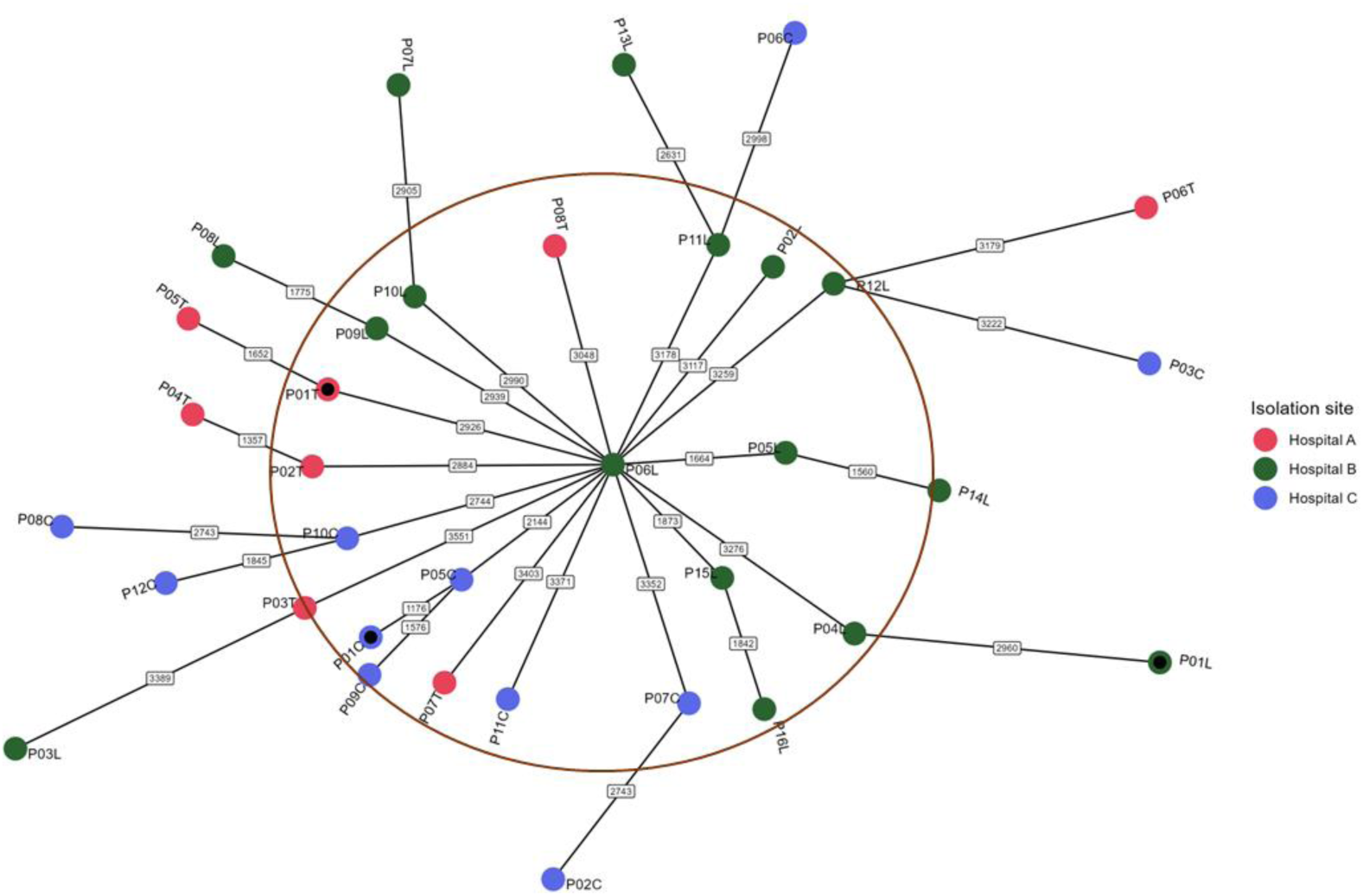
Genotyping of 35 CRPA isolates based on cgMLST schema (Tönnies et al., 2021)utilizing pyMLST (Biguenet et al., 2023) and cgmlst.org database (de Sales et al., 2020). The circles are named according to the isolates and colored according to the isolation sites. The inner black circles represent the index case. The numbers on connecting lines displayed the number of allele differences between each CRPA isolates investigated. The current cgMLST scheme has no standardized typing of *P. aeruginosa* (Biguenet et al., 2023). The major cluster involving the P06L is indicated in a circle shown in FIG 3B.

### Presence of metallo-beta-lactamases (MBLs) and extended spectrum beta-lactamase (ESBLs)

The 35 CPRA isolates obtained from three tertiary hospitals in Metro Manila from August 2022 to January 2023 revealed high genetic diversity and exhibited a non-clonal epidemic structure composed of a huge proportion of rare strains or novel STs (FIG 4). The three different isolation sites, namely Hospital A, has an 815-bed capacity, Hospital B has a 650-bed capacity, and Hospital C has a 700-bed capacity.

**FIG 4.**
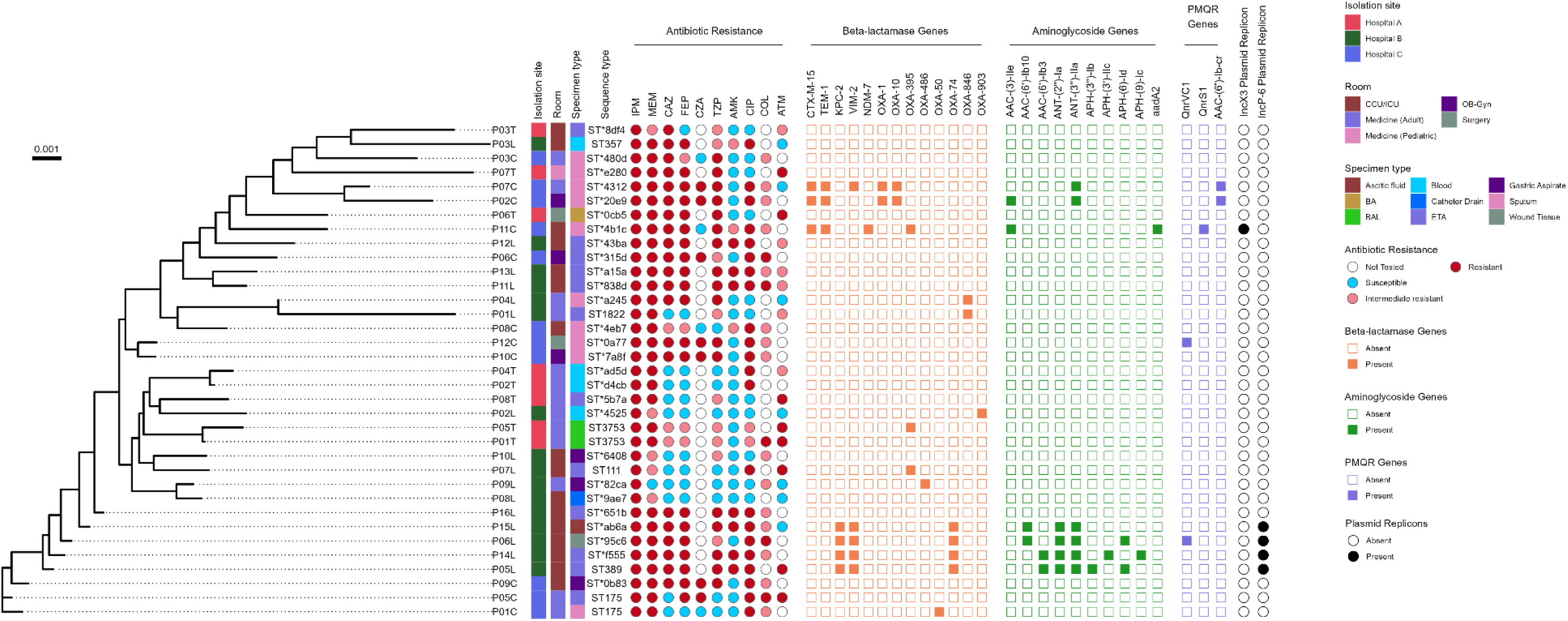
Phylogenetic SNP analysis of 35 CRPA obtained from three hospitals (Hospitals A, B And C) in Metro Manila, Philippines, from August 2022 to January 2023, the corresponding resistance profile, presence of AMR determinants, and specimen type. SNP analysis was performed using the wild-type (WT) PA01 (accession number: NC_002516.2) as the reference genome. The scale bar shows the genetic distinction between each CRPA isolates. Vertical columns demonstrate (1) isolation site, (2) patient’s room, (3) specimen type, (4) sequence type, (5) resistance phenotype: resistant (red filled circle), intermediate resistant (pink filled circle), susceptible (blue filled circle), Not Tested (empty circle), (6) resistance genotype including beta-lactamase genes: present (orange filled squares) and absent (empty squares), (7) aminoglycoside conferring resistant genes: present (green filled squares) and absent empty squares), (8) plasmid-mediated quinolone resistance (PMQR): presence (purple filled square) and absent (empty square) of acquired resistance genes, (9) predominant plasmid replicon types: presence (black filled circle) and absent (empty circle). IPM, imipenem; MEM, meropenem; CAZ, ceftazidime; FEP, cefepime; CZA, ceftazidime-avibactam; TZP, piperacillin-tazobactam; AMK, amikacin; CIP, ciprofloxacin; COL, colistin; ICU, intensive care unit; CCU, critical care unit; OB-Gyn, obstetrics and gynecology; BAL, bronchoalveolar lavage; BA, bone aspirate; ETA, endotracheal aspirate; and PMQR, plasmid-mediated quinolone resistance. Detailed results are in Supplemental File 7.

The analysis revealed that VIM-2 co-produced with KPC-2 and OXA-74 were identified in P05L, P06L, P14L, and P15L isolates obtained from patients admitted to ICU of Hospital B. In all four CRPA isolates, the KPC-2 gene was flanked by ISKpn6 And ISPay42, while the VIM-2 gene in the same isolates was flanked by Tn*As3.* P05L CRPA isolate was obtained from ETA of 73/M; P06L CRPA isolate from wound tissue of 70/M, P14L CRPA isolate was recovered from ETA of 80/F; and P15L CRPA isolates from ascitic fluid of 66/M patient (Table 3, FIG 4, FIG 5, and FIG 6). The investigations also revealed MBL VIM-2 co-produced with ESBLs TEM-1 and CTX-M-15, in P07C CRPA isolate obtained from the sputum of 70/F patient admitted to the medicine unit of Hospital C and was diagnosed as having community-acquired pneumonia. The 70/F patient was given IPM and TZP as part of her medication. The phenotypic AMR revealed resistance against IPM, MEM, CAZ, FEP, CZA, TZP, CIP and intermediate against COL. Interestingly, the IncX3 plasmid replicon type was identified from the P11C CRPA isolate that co-produced MBL NDM-7 and ESBLs TEM-1 and CTX-M-15. In addition, P11C CRPA isolate was obtained from the sputum of 25/M with meningitis admitted in the ICU of Hospital C. The 25/M patient was given the following medications: TZP, CRO, and FEP. The phenotypic AMR showed resistance against IPM, MEM, CAZ, FEP, TZP, CIP and intermediate against COL (Table 3, FIG 4, and 5).

**FIG 5.**
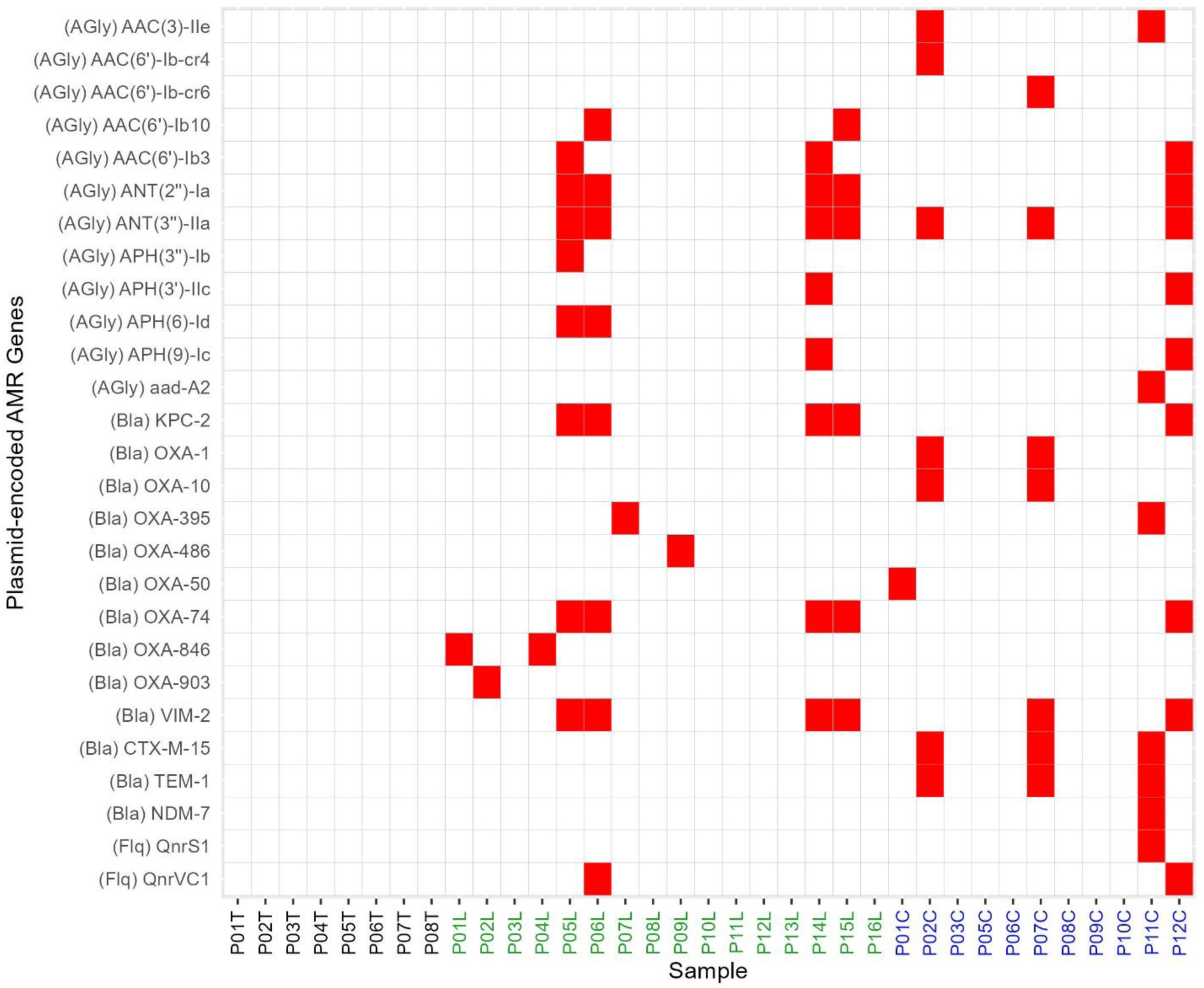
Distribution of plasmid-encoded AMR genes in 35 CRPA isolated from three tertiary hospitals in Metro Manila, Philippines, from August 2022 to January 2023. Red and white represent the presence and absence of AMR genes in the test strains. The left side denotes the category of AMR genes. Strain names are depicted at the bottom of the chart.

**FIG 6.**
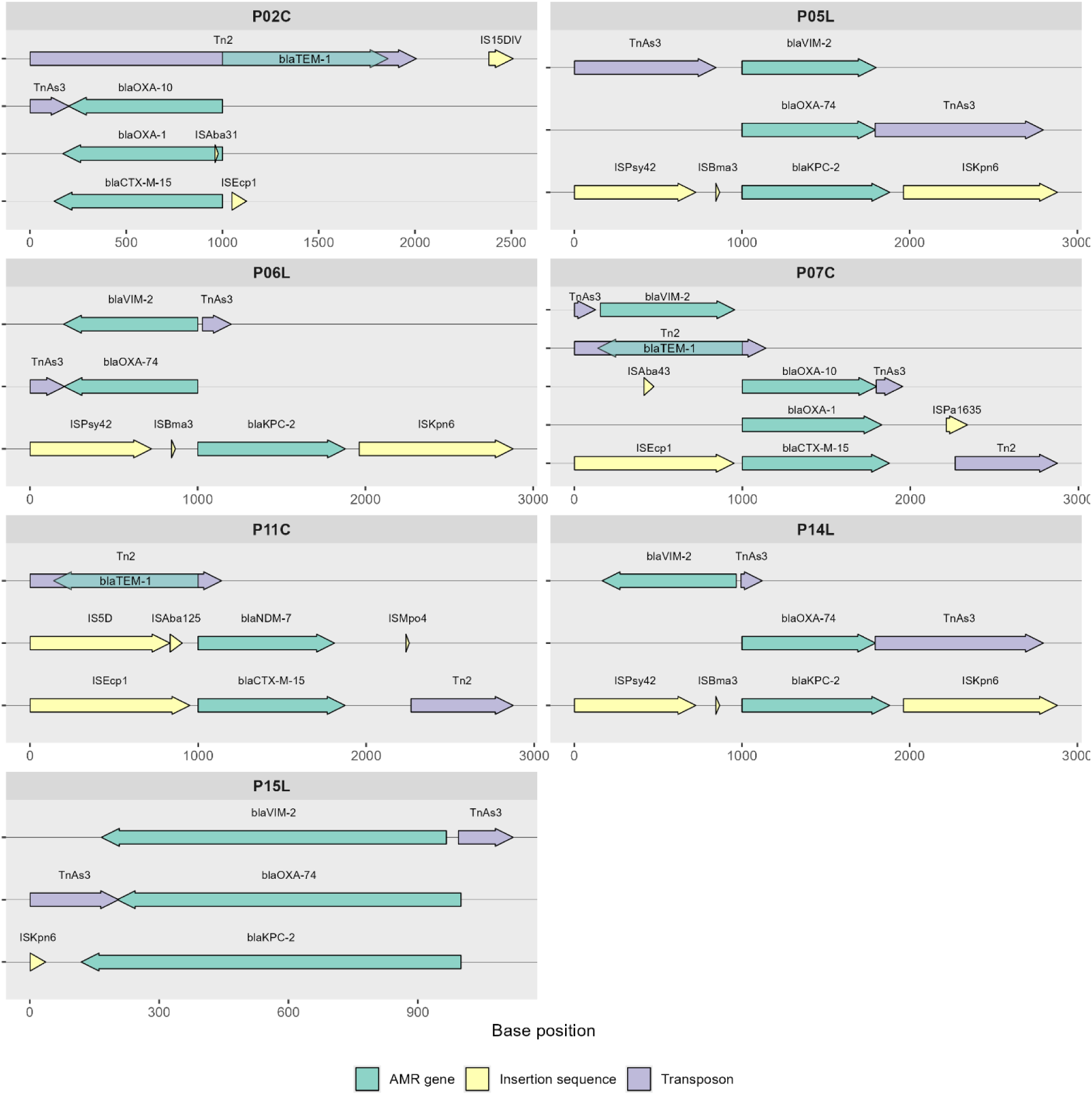
Schematic diagram of representative ARGs flanked by the MGEs in seven CRPA isolates.

**Table 3:**
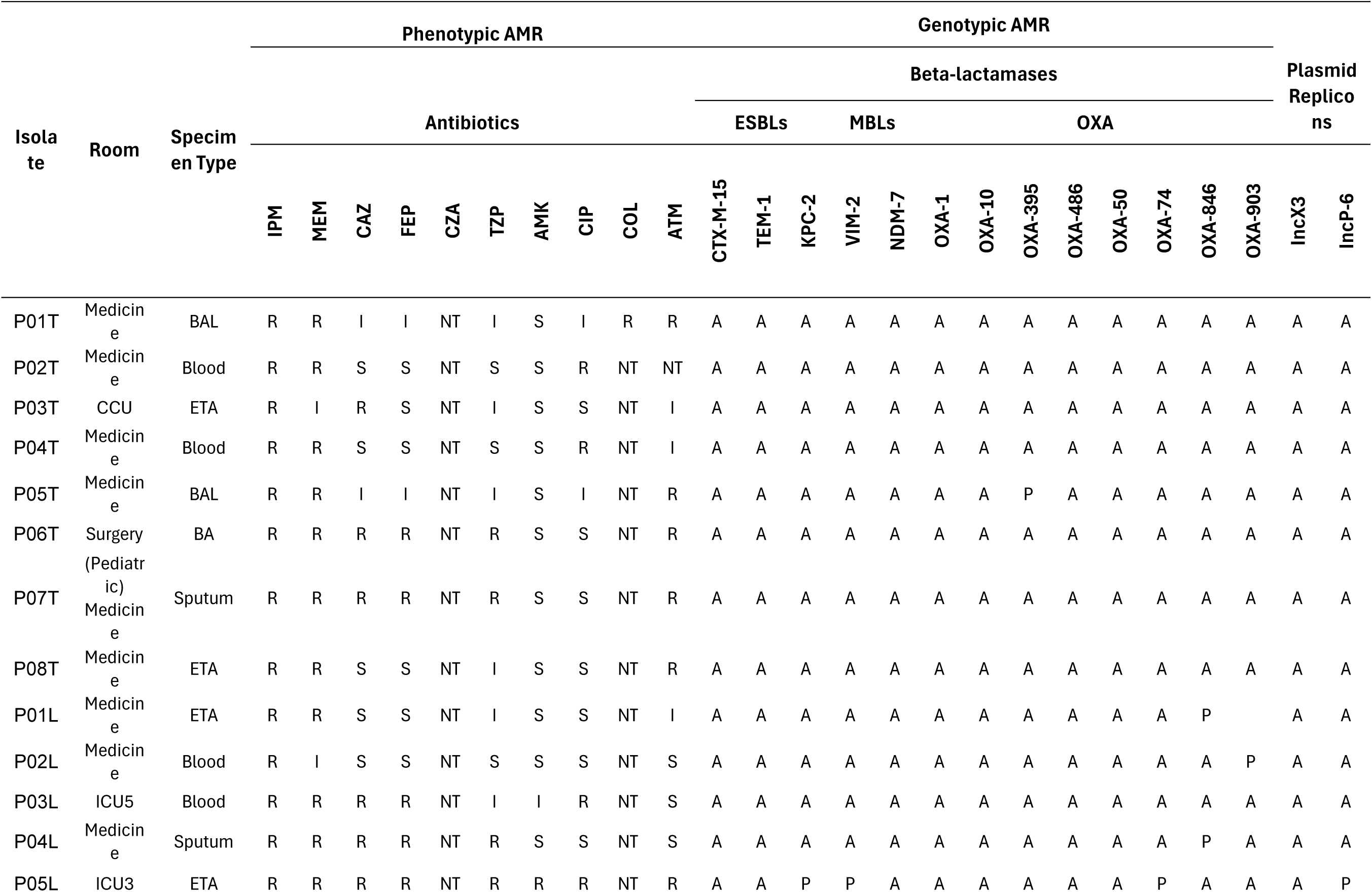
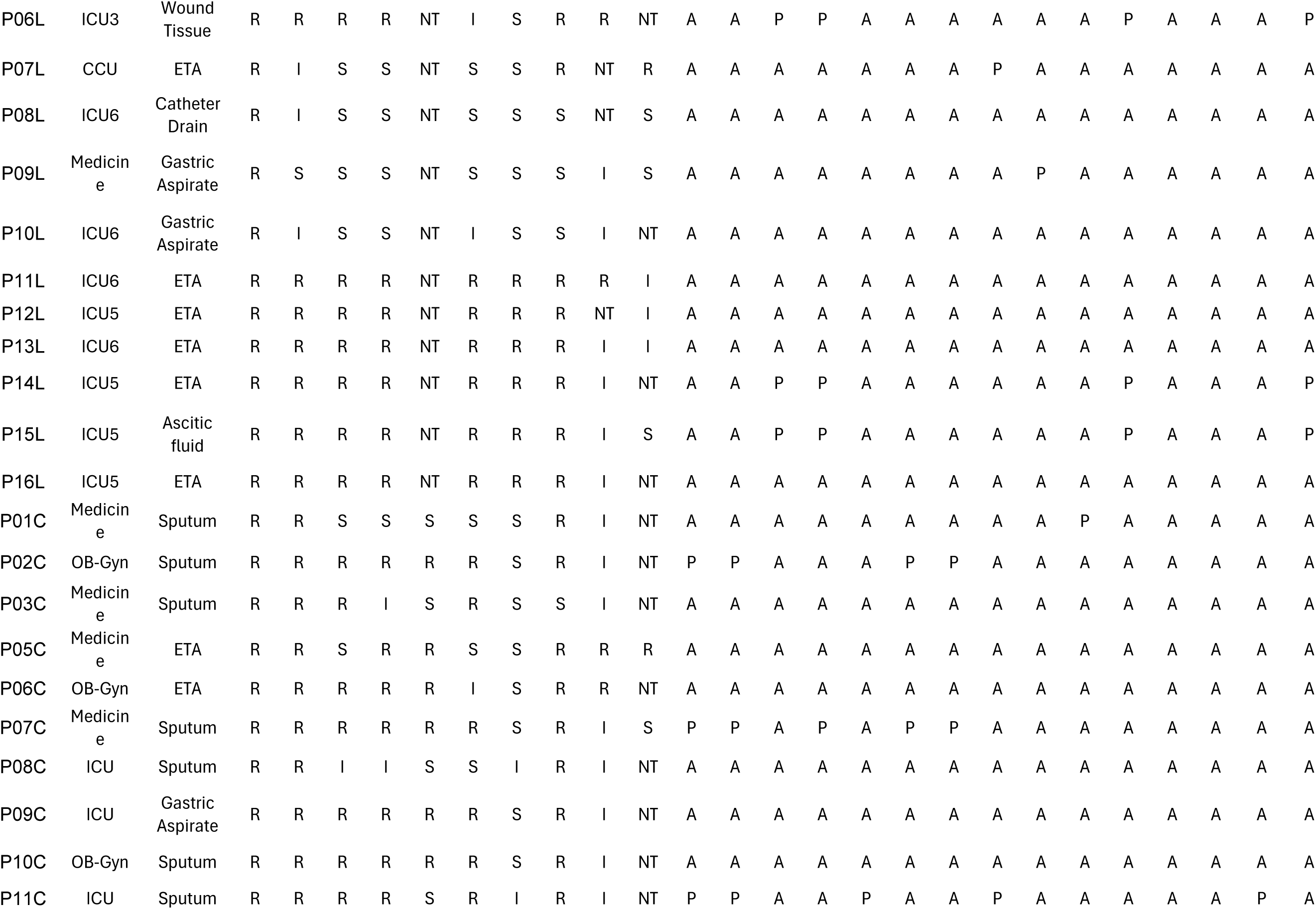
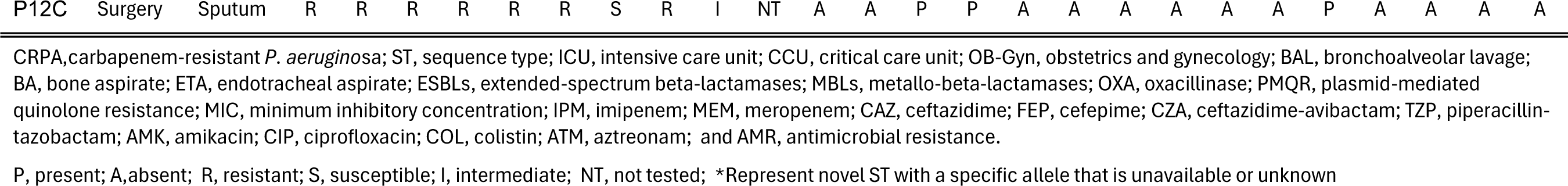
The phenotypic and the genotypic antimicrobial resistance profile of 35 CRPA isolates obtained from three hospitals.

Carbapenemases, and ESBL genes are known to be distributed by plasmids and other MGEs such as insertion sequence and transposonand speed up the spread and acquisition of ARGs (Manohar et al., 2021; Ramirez et al., 2014; Reul and et al., 2015).

#### Presence of Insertion Sequence (IS) and Transposon

Small mobile genetic elements called IS and composite transposons are essential for the transfer of antibiotic resistance genes (ARGs) among bacterial populations (Rice, 2008). Transposition mechanisms allow IS to migrate within a single cell along with resistance genes associated with them (Llaca-Díaz et al., 2013). Composite transposons, defined as areas enclosed by two identical or similar IS copies, have the ability to transport resistance genes and facilitate their transfer between distinct DNA molecules (Siguier et al., 2015). The presence of drug resistance against multiple antibiotics, the potential high-risk clones identified and the presence of ARGs, specifically carbapenemases and cephalosporinases that are flanked by MGEs pose a potential threat in healthcare settings (Partridge et al., 2018). The spread of genes that confer antibiotic resistance in bacteria is facilitated by these MGEs (Yoon & Jeong, 2021). A schematic diagram of representative ARGs and MGEs is shown in Figure 6.

The KPC-2 gene, in P05L, P06L, P14L, and P15L CRPA isolates are flanked by ISK*pn6*, ISP*ay42*, and ISB*ma3*, while VIM-2 and OXA-74 genes in the same isolates are flanked by Tn*As3.* The NDM-7 gene in CRPA isolate P11C is flanked by IS5D, ISA*ba125*, and ISM*po4*, whereas TEM-1 gene in CRPA isolates P02C, P07C, and P11C is flanked by ISE*c63*, Tn2, Tn*5393*, and IS15DIV, and CTX-M-15 in the same CRPA isolates is flanked by ISE*cp1*, and Tn2. The OXA-10 in P02C and P07C isolates are flanked by TnAs3 and ISAba43 (FIG 6).

#### Presence of IncX3 and IncP-6 Plasmid Replicon Types

Plasmids have the ability to transfer AMR genes to other bacteria, which can speed up the spread and evolution of bacterial pathogens (Teixeira et al., 2023). Antimicrobial resistance genes in *P. aeruginosa* and other Gram-negative bacteria are largely transferred by the IncP-6 and IncX3 plasmids. While the IncX3 plasmids have a narrow host range but are highly stable and have great conjugation ability, making them important targets for research and surveillance in the fight against antimicrobial resistance (Guo et al., 2022), the IncP-6 plasmids have a broad host range and can transfer and replicate in most Gram-negative bacteria (Dai et al., 2016). Plasmids within the 35 sequenced CRPA isolates were differentiated based on plasmid replicon types. The IncX3 (2.9%, n = 1/35) and IncP-6 (11.4%, n = 4/35) accounted for the prevalent plasmid replicon types detected in the CRPA isolates. P11C CRPA isolate obtained from Hospital C was positive for IncX3, whereas P05L, P06L, P14L, and P15L CRPA isolates obtained exclusively from Hospital B were positive for IncP-6 (Table 3 and FIG 4). The IncX3 plasmid replicon is the most common subgroup found to harbor NDM and KPC (Guo et al., 2022; Liakopoulos et al., 2018), contrary to the single isolate found positive for IncX3 plasmid replicon in our study.

#### Presence of plasmid-mediated quinolone resistance (PMQR) genes

Due to their ability to transmit horizontally and decreased susceptibility to fluoroquinolones, PMQR genes are important in conferring *P. aeruginosa* with resistance to these antibiotics. The fluoroquinolones ciprofloxacin and norfloxacin are less effective when modified by aac(6′)-Ib-cr, whereas the Qnr genes encode proteins that protect DNA gyrase and topoisomerase (Venkataramana et al., 2022). In this study, the PMQR genes were identified in 14.3% (n = 5/35) of the CRPA isolates investigated. The QnrS1 (2.9%, n = 1/35) was identified from P11C CRPA isolate obtained from Hospital C, while the QnrVC1 was identified from (5.7%, n = 2/35) P06L CRPA isolate obtained from Hospital B and P12C isolate obtained from Hospital C. Moreover, the ciprofloxacin and aminoglycoside modifying enzyme (aac-(6’)-Ib-cr) were identified from P02C and P07C isolates obtained from Hospital C, accounting to 2.9%, (n = 1/35). Of the 62.9% (n = 22/35) CRPA isolates shown to be phenotypically CIP resistant, only 22.7% (n = 5/22) harbored the PMQR genes (Table 3 and FIG 4 and 5). Additionally, this study looked at mutations in *gyr*A and *par*E that could also contribute to the CIP resistance (Supplemental File 6B).

#### The presence of mutation in the Quinolone Resistance-Determining regions (QRDR) - *gyrA* and *parE* genes

Resistant to CIP in *P. aeruginosa* has been associated with mutations in the QRDR of the *gyr*A and *parE* genes. It has been demonstrated that high-level resistance to CIP in *P. aeruginosa* isolates is encoded by mutations in both the *gyr*A and *par*E genes. Furthermore, mutations in the *par*E gene, specifically when they coexist with mutations in *gyr*A or *par*C, are a contributing factor to high-level CIP resistance (Arabameri et al., 2021). The combination of these mutations results in structural changes to DNA gyrase, which lowers the enzyme’s susceptibility to antibiotics and promotes drug resistance (Arabameri et al., 2021).

SNPs *T83I* was identified in the *gyrA* gene A473V was identified in the *par*E gene in 31.4% (n = 11/35) and 5.7% (n = 2/35) CRPA isolates investigated. T83I and A473V mutations have been known to cause resistance to fluoroquinolones. The CRPA isolates that harbored *gyrA* mutations (T83I) are isolates: P02T, P04T, P05L, P06L, P14L, P15L, P01C, P02C, P05C, P07C and P11C, while P02T and P04T isolates obtained from Hospital A harbored *parE* mutations (A473V). All CRPA isolates that exhibited both or either of the *par*E and *Gyr*A mutations and at least one of the PMQR genes demonstrated resistance against CIP. However, 45.4% (n = 10/22) of CRPA isolates that displayed phenotypic resistance against CIP did not possess *gyr*A and *par*E mutations, including PMQR gene expression ( Supplemental File 6B and 7).

#### Presence of aminoglycoside conferring resistance genes

Genes mediating aminoglycoside resistance were also identified from 20.0% (n = 7/35) of CRPA isolates investigated exclusively recovered from Hospital B and C. ANT-(3’’)-IIa has the highest proportion among the identified aminoglycoside conferring resistance genes, 17.1% (n = 6/35). ANT(2’’)-Ia, APH(6)-Id, APH(3’’)-Ib, APH(3’)-IIc, AAC(3)-IIe, AAC(6’)-Ib10, And AAC(6’)-Ib3, were also detected from both Hospital B and C isolates and each gene accounting to 5.7% (n = 2/35). Also present are APH(3’’)-Ib, APH(3’)-IIc, APH(9)-Ic, and aadA2 each gene accounting to 2.9% (n = 1/35). Of these, 43.0% (n = 3/7) were phenotypically expressed. Moreover, the resistance determinants for the observed AMK resistance were not detected in 57.1% (n = 4/7) of the CRPA isolates investigated (FIG 4 and 5 and Supplemental File 7).

#### Incidence of OprD mutations

Resistance in CRPA isolates that do not produce carbapenemases could be mediated by a loss of porin D (OprD) and/or overexpression of efflux pumps (such as MexAB-OprM) (Amsalu et al., 2020; Arzanlou et al., 2017; Gashaw et al., 2024; Nieto-Saucedo et al., 2023). OprD is an outer membrane porin in *P. aeruginosa* that facilitates the uptake of basic amino acids and imipenem (Alcock et al., 2023). However, mutations in the OprD that will correlate with carbapenem resistance, such as Gln143X, were not detected in all CRPA isolates investigated (Supplemental File 6A).

## DISCUSSION

*P. aeruginosa* that produce beta-lactamases specifically carbapenemases is a global problem. The assessment was performed utilizing an in-depth analysis of the whole genome sequence of 35 non-duplicate CRPA isolates obtained from patients admitted from three hospitals collected over the period of six months. Despite measures implemented in the isolation sites to combat AMR through antimicrobial stewardship, surveillance, and understanding of AMR, the prevalence of AMR remains a significant challenge in patient care, particularly in Hospitals B and C, with a high proportion of CRPA isolates carrying acquired AMR genes, and a moderate to high level of resistance against the last line of antibiotics. Four of the so-called “big five carbapenemases,” namely NDM, VIM, KPC, And OXA, were identified in this study.

This article first reported on the co-production of KPC-2, VIM-2, and OXA-74 represented by potential high-risk clones *P. aeruginosa* ST389, ST*95c6, ST*f555, and ST*ab6a, and in the Philippines. In hospital settings, epidemic outbreaks of *P. aeruginosa* MDR/XDR high-risk clones are usually associated with ST111 And ST175 (Cabot et al., 2012; Horcajada et al., 2019; Kocsis et al., 2021; Oliver et al., 2015); however, this was not the outcome in our investigation of the 35 CRPA isolates obtained from the three hospitals. Hospitals B and C displayed a high proportion of CRPA isolates carrying acquired AMR genes, specifically carbapenemases such as VIM-2 and co-produced with KPC-2 and OXA-74, whereas Hospital C had a large proportion of CRPA isolates carrying ESBLs such as CTX-M-15 and TEM-1. In contrast, in the Department of Health’s (DOH) Antimicrobial Resistance Surveillance Reference Laboratory (ARSRL) annual report, VIM, NDM, and ESBLs were not observed; instead IMP-4 was detected in one *P. aeruginosa* isolate obtained from an in-line catheter (Department of Health - RITM, 2022). In addition to the rapid dissemination and acquisition through horizontal gene transfer, NDM, VIM, KPC, and some OXA variants are plasmid-borne carbapenemase-encoding genes that present challenges in the treatment of bacterial infections. These genes have the capacity to hydrolyze nearly all beta-lactam antibiotics, including carbapenems (Leungtongkam et al., 2018; Manohar et al., 2021; Murray et al., 2022; Reyes et al., 2023; US Department of Health and Human Services; CDC., 2019).

In our report, KPC-2, VIM-2, OXA-74, NDM-7 and ESBLs were flanked by MGEs that will add burden in the mitigation of spread and acquisition through horizontal gene transfer. In addition, one of five (1/5) CRPA isolates that were identified as positive for KPC-2 and VIM-2 was found resistant to CZA. Further surveillance and investigation are suggested since it was studied that when it comes to isolates that produce KPC, CZA is highly active while remaining inactive against pathogens that produce MBLs (Horcajada et al., 2019). However, the same isolate was also positive for MBL VIM-2 that added burden to treatment management. A study reported KPC-2 genes located between IS*Kpn6*-like and IS*Kpn8*-like MGEs (Hu et al., 2021). However, in our study, KPC-2 is located between IS*Psy42* and IS*Kpn6,* VIM-2 is flanked by TnAs3, while NDM-7 is surrounded by IS5D and IS*Mpo4*. These can facilitate the movement of resistance genes between different DNA molecules (Rice, 2008).

A multinational study identified the differences in the prevalence and types of carbapenemases harbored by CRPA across different regions and found that carbapenemases were rare in CRPA isolates in the U.S. but common in isolates in other regions such as South And Central America, particularly KPC-2 and VIM-2 (Reyes et al., 2023). Another study reported that CRPA ranges from 10 to 30% in countries such as Australia and North America to more than 50% in Saudi Arabia, Brazil, Iran, Poland, Peru, Greece, Russia, and Costa Rica, where resistance rates are high enough to be concerning for public health (Halat & Moubareck, 2022; Hong et al., 2015b). Reports confirmed the geographic differences in carbapenemase genotypes, such as the U.S. epidemic was linked to KPC (class A in the Ambler classification) (Bush & Jacoby, 2010), the European epidemic to VIM (class B) and OXA-48-like (class D), and the Asian epidemic to NDM and IMP (class B) (Suzuki et al., 2019). The NDM type is epidemic in clinical settings in the Philippines. In fact, a study on beta-lactam-resistant Gram-negative bacilli clinical isolates in a Philippine tertiary care hospital co-produced NDM-1, TEM-5, CTX-M-117, IMP-4, OXA-1, and OXA-72 in one *P. aeruginosa* isolate, while another *P. aeruginosa* isolate co-harboring NDM-1, IMP-1, OXA-1, and OXA-72 (Abordo et al., 2023). Similarly, we also identified in our study, P11C a CRPA isolate carrying NDM-7 that co-produced CTX-M-15, TEM-2, and OXA-395 and is represented by novel ST*4b1c. This isolate was positive for the IncX3 type plasmid replicon and the ARGs were surrounded and flanked by MGEs making the P11C CRPAS a potential high-risk clone. The P11C CRPA isolate from Hospital C was obtained from the sputum of a 26-year-old male ICU patient who was treated with piperacillin-tazobactam, ceftriaxone, and cefepime. Furthermore, another investigation underscored the global clinical implications of CRPA, noting that its genetic and epidemiological attributes are poorly understood (Chen et al., 2018), and there is a need for further research to understand their genetic and epidemiological attributes. In this article, we elucidated the genetic and epidemiologic characteristics of the 35 CRPA strains, specifically the co-production of VIM-2 and KPC-2 in P05L, P06L, P14L, and P15L CRPA strains obtained from ICU patients in Hospital B. These CRPA strains are both IPM and MEM-resistant. In other published studies, ST175 is frequently observed to produce VIM-2, whereas ST111 can produce KPC-2 carbapenemase (Oliver et al., 2015); however, this was not the case in our investigation because ST175 and ST111 did not contain any MBLs and ESBLs, instead, the novel STs were the clones responsible for the co-production of VIM-2, KPC-2, and OXA-74. The genetic heterogeneity of 35 CRPA isolates investigated in this study contrasts with the clonal dominance of high-risk clones in other studies (Cabot et al., 2012; Oliver et al., 2015; Pollini et al., 2013).However, the limited sample size and short duration of sample collection can be a factor.

The 35 CPRA isolates obtained from three tertiary hospitals in Metro Manila from August 2022 to January 2023 exhibited high phenotypic and genetic diversity (FIG 6). The presence of multiple drug resistance CRPA strains and multiple antibiotic-resistance genes that are flanked by mobile genetic elements (MGE) (FIG 6) pose a potential threat in healthcare settings. A more stringent protocol needs to be in place, particularly for the sites that showed a high proportion of AMR and a high level of resistance against the last line of treatment.

## CONCLUSIONS

This research presented an insight into the current genetic characteristics of 35 CRPA isolates obtained from August 2022 to January 2023 from the three tertiary hospitals in Metro Manila, Philippines, highlighting the first identification of the CRPA harboring plasmid-borne VIM-2, and co-produced with KPC-2, And OXA-74 flanked by MGEs. Additionally, the persistent identification of NDM in the Philippine’s clinical setting remains a major global public health concern. Control strategies should be strongly implemented, along with discovering inhibitors targeting carbapenemase-producing *P. aeruginosa*. The genetic and epidemiologic characterization of clinical CRPA isolates, in addition to surveillance, are of the utmost importance in order to enhance the outcomes of infection control measures designed to mitigate the incidence and spread of CRPA infection by providing current information.

## MATERIALS AND METHODS

### Isolation and Collection of *P. aeruginosa*

From August 2022 to January 2023, 40 CRPA isolates were obtained from the three tertiary hospitals in Metro Manila, Philippines. The Vitek®2 system (bioMérieux) and/or BD Phoenix™ M50 instrument was used for species identification, and the strains that showed imipenem and/or meropenem resistance with MIC breakpoints of ≥8.0 mg/L as the result of antibiotic susceptibility using the Vitek®2 system (bioMérieux) and /or BD Phoenix™ M50 instrument, were collected. *P. aeruginosa* Strains that demonstrated MIC breakpoints of ≥8.0 mg/L to carbapenem (imipenem and or meropenem) were designated as CRPA and were included in the study provided that the source CRPA isolates are the following: lower respiratory tract, wound discharge, blood, and other sterile body fluids. Other sources, such as stool, throat, And nasal swabs, were excluded from the collection. Out of 40 CRPA isolates, 36 were successfully sub-cultured, and pure colonies were isolated for DNA extraction. Thirty-six CRPA strains were subsequently subjected to WGS, but one sample was removed due to low-quality sequence reads, and 35 were included in downstream analysis.

### Antimicrobial susceptibility test

The antibiotic susceptibility test was performed using the GN N261 CARD of Vitek®2 system (bioMérieux) for the following Antibiotics: IPM, MEM, FEP, CAZ, CIP, TZP, And AMK. In addition, the MIC for COL and CZA was determined using the EURGNCOL plate of the Sensititre system™ (Thermo Fisher Scientific™) via the commercial broth microdilution kit And the standard broth microdilution (BMD), performed according to the CLSI CLSI M100 ED32:2022 guideline (CLSI, 2022). Kirby Bauer technique for the determination of antimicrobial resistance against ATM was used. The Antimicrobial susceptibility of the strains was determined according to the guidelines And breakpoint of the Clinical Laboratory Standard Institute (CLSI) M100-ED32:2022, with *P. aeruginosa* ATCC 27853 used as the quality control strains (CLSI, 2022).

### DNA extraction

DNA was extracted using DNeasy® UltracleAN® Microbial Kit (Qiagen^TM^, Germantown, MD, USA) from cells grown overnight at 37 °C in Tryptic Soy Broth (TSB) following the manufacturer’s protocol with slight modifications in the speed and time set for centrifugation. DNA concentrations were measured on Qubit using Invitrogen Qubit^TM^ 4 Fluorometer. For the quality of the DNA, it was performed using FLUOstar® Omega.

### Whole-genome sequencing and analysis

The DNA extracts of 36 CRPA clinical isolates underwent sequencing in SA Pathology (Adelaide, Australia) using the Illumina NextSeq 550 platform employing 150-base pair-end reads sequencing. The Amplicon Library was prepared using the Nextera XT DNA Library Preparation kit (Illumina Inc., USA) following the manufacturer’s protocol.

### Data Processing and Analytics

The raw sequence data were uploaded onto an in-house bioinformatics pipeline, called EpiTomas v1.0.0 (https://github.com/abulenciamiguel/EpiTomas). EpiTomas offers an automated workflow for quality control evaluation, reference-based assembly, de novo assembly, and genome characterization (FIG 7).

**FIG 7.**
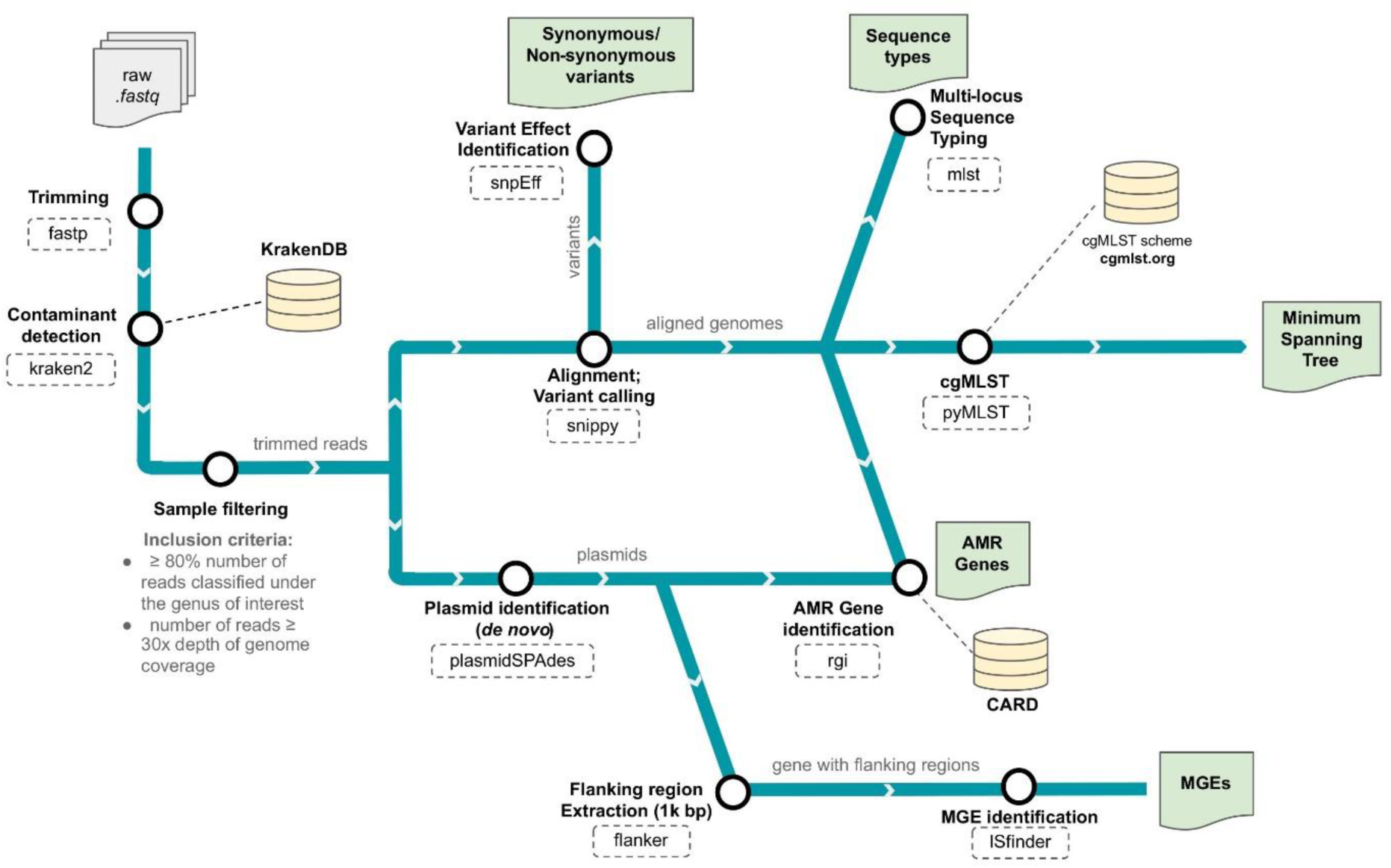
The in-house bioinformatics pipeline was used in the analysis. Available at https://github.com/abulenciamiguel/EpiTomas

### Raw read quality control

The raw sequences underwent read trimming and quality assessment using Fastp (Chen et al., 2018). Thirty-five (35) CRPA isolates met the quality control standards, having sequence read with a Phred score of 30 (Q30), ensuring a high level of accuracy, and 80% of the reads that passed the filter belonged to *Pseudomonas* genus using Kraken2 Minikraken (Wood et al., 2019) database, with the number of reads providing at least 30x read coverage, ensuring comprehensive coverage of the genome and the genome must have a GC content ranging from 65 to 67%.

### Sequence alignment against PA01 RefGenome and varint calling

After pre-processing, reads underwent alignment against PA01 NC_002516.2 as the reference genome using snippy, and subsequently determined the alignment coverage using mosdepth (Pedersen & Quinlan, 2018). Additionally, the predicted effect and annotation of the called variants were performed using SnpEff (Cingolani et al., 2012). The PAO1, as the chosen reference genome, was completely sequenced by the year 2000, and its genome is relatively large (5.5–7 Mbp) in comparison to other bacteria that have been sequenced (Stover et al., 2000). Furthermore, plasmid contigs of the 35 CRPA strains were assembled from the trimmed fastq files using plasmidSPAdes (Antipov et al., 2016).

### AMR genes And flanking mobile genetic elements detection

The assembled genomes and the plasmid contigs undergo further analysis, such as identification and characterization of AMR genes using the Resistance Gene Identifier (RGI) with the Comprehensive Antibiotic Resistance Database (CARD) as a reference (Alcock et al., 2020). Mobile genetic elements flanking 1,000 bp up- and downstream the selected seven AMR genes (i.e., *bla*VIM-2, *bla*KPC-2, *blaND*M-7, *bla*TEM-1, *bla*CTX-M-15, *bla*OXA-74, *bla*OXA-1, and *bla*OXA-10) in plasmid contigs were identified using ISfinder webtool (Siguier et al., 2006).

### Traditional MLST and cgMLST analyses

Genotyping was performed using MLST for 35 CRPA isolates obtained from the three tertiary hospitals in Metro Manila, Philippines. *P. aeruginosa* MLST profiles were predicted using Pasteur nomenclature in the Pathogenwatch (https://cgps.gitbook.io/pathogenwatch/technical-descriptions/typing-methods/mlst) and PubMLST database (Curran et al., 2004; Jolley et al., 2018; Jolley & Maiden, 2010). Pathogenwatch is a free, accessible, real-time microbial genome analysis platform that provides detailed and integrated genomic and epidemiological data. PubMLST provides Pathogenwatch’s schemes, and an in-house search tool is used to ensure accurate MLST and cgMLST assignments (Sánchez-Busó et al., 2021). MLST is based on a community-agreed list of seven gene loci found in all species strains, with a code assigned to each allele (Curran et al., 2004; Jolley et al., 2018; Jolley & Maiden, 2010).

Additionally, cgMLST schema (Tönnies et al., 2021) a high-resolution typing was conducted using pyMLST (Biguenet et al., 2023) and the cgMLST.org database (de Sales et al., 2020) because of the ubiquitous characteristics of the 35 CRPA isolates and the involvement of multiple sequence types both known and new STs revealed by the traditional MLST. pyMLST is a Python tool that uses a broader selection of genes from the complete or core genome for bacterial phylogeny and typing (Biguenet et al., 2023). The strain’s sequence type (ST) is determined by the combination of the several alleles that each distinct sequence corresponds to (Biguenet et al., 2023). pyMLST allows for the iterative expansion of strain collections for comparison by storing MLST profiles and allele sequences in a local SQLite database (Biguenet et al., 2023; Tönnies et al., 2021). Using BLAST-Like Alignment Tool (BLAT). PyMLST’s method aligns the first allele of each gene in the cg/wgMLST schema to the target bacterial genome of interest Biguenet et al., 2023). MAFFT is a tool used to align incomplete genes in the cg/wgMLST schema, allowing precise data analysis and SQLite database storage (Biguenet et al., 2023). With PyMLST, users can generate schemas from their own data or download pre-existing schemas straight from sites like https://www.cgmlst.org/ncs. In this case, the cgMLST.org database was used. The 1,316 whole genomes of *P. aeruginosa* that were obtained from the NCBI were used to create the cgMLST.org database, which can anaalyze 1,265,835 alleles in the nomenclature server and 3,867 locus counts (de Sales et al., 2020; Tönnies et al., 2021).

### Phylogenetic Analysis

The minimum spanning tree was constructed using GrapeTree’s MSTreeV2 (72) algorithm to assess the *P. aeruginosa* clonality based on allele differences and determine the epidemiological landscape. In addition, IQ-TREE2 ( Minh et al., 2020) was used to construct the maximum likelihood tree with 1000 bootstrap replicates to assess *P. aeruginosa* genetic diversity based on single nucleotide mutation.

## Data availability

Reads from all sequence samples (fastq files) are available under the BioProject accession PRJNA1045071 and were made public in July 2025.

## Ethics declarations

Ethical permission was approved by the Ethics Committee of the University of Santo Tomas Hospital on August 2022 under Protocol Reference No.: REC-2022-04-061-IS And the Institution’s approval for the two hospitals.

## Funding

No funding sources

## Conflict of interest

No conflict of interest.

## SUPPLEMENTAL MATERIALS

Excel File for Supplemental Files 1 to 7 .

## ACKNOWLEDGMENT

Isolates were provided by the Microbiology Department of the University of Santo Tomas Hospital, St. Luke’s Medical Center, Quezon City, and Chinese General Hospital and Medical Center, Manila. Assistance from Ms. Florelei Ordañez Alva, RMT; Mr. Miguel Alpez, RMT; Mr. Russell Gambito Panem, RMT; and Ms. Maria Eliza G. Apo, RMT.

Assistance was provided by Ms. Therris Mae Mamerto, MSc, Mr. Rinnel Bonifacio, Mr. Cleve Vergara, and Mr. Adriane Villavieja of the CMDR – UST laboratory.

Coordination from A/Prof. Matthew Sykes, Ph.D. of the University of South Australia, Prof. Edilberto P. Manahan, Ph.D., Prof. Agnes Castillo, Ph.D., and Prof. Dean Aleth Therese Dacanay, Ph.D. of the Faculty of Pharmacy, University of Santo Tomas.

The authors would also like to acknowledge Aldrin V. Imbag and Francisco Gerardo M. Polotan for providing the compute resources utilized in this study.

## Notes

### Competing Interest Statement

The authors have declared no competing interest.

### Funding Statement

This study did not receive any funding.

### Author Declarations

Ethical permission was approved by the Ethics Committee of the University of Santo Tomas Hospital on August 2022 under Protocol Reference No.: REC-2022-04-061-IS and the institution approval for the Chinese General Hospital and Medical Center and St. Luke's Medical Center.

